# Clonal diversity determines persistence of SARS-CoV-2 epitope-specific T cell response

**DOI:** 10.1101/2022.04.18.22273961

**Authors:** Ksenia V. Zornikova, Alexandra Khmelevskaya, Savely A. Sheetikov, Dmitry O. Kiryukhin, Olga V. Shcherbakova, Aleksei Titov, Ivan V. Zvyagin, Grigory A. Efimov

## Abstract

T cells play a pivotal role in reducing disease severity during SARS-CoV-2 infection and formation of long-term immune memory. We studied 50 COVID-19 convalescent patients and found that T cell response was induced more frequently and persisted longer than circulating antibodies. To identify epitopes that give rise to long-lived T cell memory, we performed *ex vivo* T cell expansion, MHC-tetramer cell-sorting, and high-throughput sequencing. We identified 756 clonotypes specific to nine known CD8^+^ T cell receptor (TCR) epitopes. Some epitopes were recognized by highly similar public clonotypes with restricted variable and joining segment usage. Receptors for other epitopes were extremely diverse, suggesting alternative modes of recognition. We also tracked persistence of epitope-specific response and individual clonotypes for a median of eight months after infection. The number of recognized epitopes per patient and quantity of epitope-specific clonotypes decreased over time, but the studied epitopes were characterized by uneven decline in the number of specific T cells. Epitopes with more clonally diverse TCR repertoires induced more pronounced and durable responses. In contrast, the abundance of specific clonotypes in peripheral circulation had no influence on their persistence. Our study demonstrates the durability of SARS-CoV-2-specific CD8^+^ memory, and offers important implications for vaccine design.

## Introduction

The ongoing pandemic of Coronavirus disease 2019 (COVID-19), caused by the novel severe acute respiratory syndrome coronavirus 2 (SARS-CoV-2), has resulted in considerable morbidity and mortality worldwide. It is well established that recovery from COVID-19 is mediated not only by the production of neutralizing antibodies but also the development of a T cell response to SARS-CoV-2 antigens (Grifoni et al., 2020; Le Bert et al., 2020; Braun et al., 2020; Peng et al., 2020).

The levels of circulating binding and neutralizing antibodies are highly variable between individuals, but in most patients, humoral response decays over the first 6–8 months post-infection (Dan et al., 2021; Sherina et al., 2021, Wheatley et al. 2021). It has been shown that SARS-CoV-2-specific memory B cells (Sherina et al., 2021; Byazrova et al., 2021a; Byazrova et al., 2021b; Sokal et al., 2021; Marcote et al., 2021) and T cells are maintained in COVID-19 convalescent patients at least for 6–8 months (Dan et al., 2021; Sherina et al., 2021; Zuo et al., 2020; Bilich et al., 2021), and the latest reports further extend this margin to up to 10–15 months (Jung et al., 2021, Marcote et al., 2021). Although the numbers of circulating T cells decrease over time (Wheatley et al., 2021; Marcotte et al., 2021), these cells retain the ability to secrete cytokines (Kang et al., 2021) and to expand (Rodda et al., 2021) upon re-stimulation with the appropriate antigen.

SARS-CoV-2-specific T-cell responses are more prevalent than seropositivity, and can be detected in the majority of the patients who have not developed specific antibodies (Schwarzkopf et al., 2021; Bonifacius et al., 2021). This makes the assessment of specific T cell reactivity an accurate indicator of past infection (Tan et al., 2021; Titov et al., 2021). In some cases, pre-existing T cell responses might even prevent the development of full-blown disease (Swadling et al., 2021; Titov et al., 2021).

Mounting evidence points to the clinical importance of T cells in COVID-19. Specific CD4^+^ and CD8^+^ T cell responses contribute to reduced severity of disease (Rydyznski Moderbacher et al., 2020; Zhou et al., 2020; Liao et al., 2020). Early induction of a functional SARS-CoV-2-specific T cell response is associated with rapid viral clearance and milder disease (Tan et al., 2021b). On the contrary, T cell anergy is connected with a poor prognosis (Renner et al., 2020). Reduced diversity of T cell response is characteristic of patients with COVID-19 pneumonia (Chang et al., 2021), while decreased numbers of CD8^+^ T cells indicate a high probability of developing severe disease and even death (Wang et al., 2020).

There is evidence that T cells can successfully clear the virus even in the absence of antibodies. Patients with humoral immunodeficiency or depleted B cells successfully recover from COVID-19 (Jones et al., 2021; Asplund Högelin et al., 2021; Hughes et al., 2021). Moreover, in a large prospective study, we have recently shown that a subgroup of patients lacking a humoral response to SARS-CoV-2 was partly protected from infection by T cells (Molodtsov et al., 2021). A beneficial role of T cells has also been shown in mice: in the absence of antibodies, T cells were able to eliminate SARS-CoV infection, and immunization with a single immunodominant CD8^+^ T cell epitope protected against lethal disease (Zhao et al., 2010; Channappanavar et al., 2014).

We and others have identified multiple CD8^+^ and CD4^+^ immunogenic SARS-CoV-2 epitopes (Ferretti et al., 2020; Nelde et al., 2020; Shomuradova et al., 2020; Gangaev et al., 2021; Kared et al., 2021; Saini et al., 2021). Recently, we systematically characterized the immunogenicity of a panel of unique SARS-CoV-2 epitopes presented by the most common HLA alleles (Titov et al., 2021). A number of publications have reported sequences of T cell receptors (TCRs) recognizing some immunodominant epitopes of SARS-CoV-2 (Shomuradova et al., 2020; Chaurasia et al., 2021; Minervina et al., 2021; Snyder et al., 2020). It was demonstrated that TCRs specific to some epitopes are public (Shomuradova et al., 2020; Simnica et al., 2021; Nguyen et al., 2021) and have a high degree of mutual similarity, which is sometimes accompanied by strong biases in variable (V) and joining (J) gene usage. According to the structural data, the latter can be explained by germline-based epitope recognition (Wu et al., 2022). The notion of SARS-CoV-2-specific TCRs could be used to identify patients exposed to this virus by analyzing individual TCR repertoires (Snyder et al., 2020; Shoukat et al., 2021; Mayer-Blackwell et al., 2021).

Recurrent mutations in the Spike (S) protein have given rise to variants of concern, including the current omicron variant, which are less susceptible to neutralization by antibodies (Cele et al., 2021). At the same time, the large numbers and variability of T cell epitopes recognized in different individuals make T cell response more resilient against immune evasion. Several non-synonymous mutations in known T cell epitopes have been reported, resulting in diminished or abrogated MHC binding or T cell activation (Agerer et al., 2021; Wu et al., 2021). However, these mutations only seldom get fixed into variants of concern, and this is probably because variants escaping presentation by one HLA allele often become binders to other alleles (Nersisyan et al., 2021).

One of the key remaining questions is what factors influence the longevity of SARS-CoV-2-specific T cell response. Careful analysis of T cell response characteristics at the level of individual epitopes and clones might unravel factors influencing the formation and persistence of long-lived T cell memory. To this end, we have measured humoral and T cell responses to SARS-CoV-2 in paired blood samples of 50 COVID-19 convalescent patients (CP) shortly after infection and at a median of 8 months after infection. Moreover, for a subgroup of 26 CP, we characterized the frequency of memory T cells, their clonal structure, and the TCR repertoire of the T cell immune response to a panel of eight CD8^+^ epitopes. We found that T cell response was induced more often than seropositivity and persisted in a larger group of patients. Nevertheless, cellular immunity also diminished over time, manifesting as a decrease in the number of recognized antigens and epitopes, reduced frequency of specific T cells in circulation, and a reduced number of epitope-specific clonotypes. Still, most epitopes were recognized for up to 10 months post-infection.

We have described a total of 756 epitope-specific T cell clonotypes and demonstrated that the response to some of these epitopes was mediated by public TCRs and TCRs with highly similar CDR3β amino acid sequences, whereas others were recognized by highly diverse receptors. Most epitope-specific clonotypes were present at levels below the limit of detection in peripheral blood. The most important factor influencing the dominance of response to a particular epitope and its persistence was the number of specific clonotypes detected after infection, while the average size of the clonotypes did not have a significant impact. In the face of neutralizing antibody escape by emerging variants, cellular immunity has growing importance in terms of reducing the severity of COVID-19 infection. So far, most widely-used COVID-19 vaccines are based on the S protein; in contrast, of the nine epitopes designated as immunodominant in this work, only three are S-derived. This study provides a rationale for including immunodominant T cell epitopes in the new generation of vaccines to ensure the formation of long-term T cell memory.

## Results

### Higher incidence and superior persistence of cellular over humoral response

To study the dynamics of adaptive immune response to SARS-CoV-2, we recruited a cohort of 50 convalescent donors (CDs) who had asymptomatic (n = 2), mild (n = 31), or moderate to severe (n = 17) COVID-19 according to the classification used by the U.S. National Institutes of Health. None of the patients required treatment in the intensive care unit, oxygen supplementation, or invasive ventilation support. The cohort included 27 females and 23 males aged 17–64 years (median = 36). Peripheral blood was collected at two time-points: between 17 and 72 days (median = 35) after disease onset (TP1), and between 180 and 292 days (median = 242) after disease onset (TP2). None of the donors had been vaccinated against COVID-19 or had confirmed reinfection between samplings. A control group of healthy donors (HD) included 19 individuals. Fourteen samples from healthy donors cryopreserved no later than August 2019 were obtained from the biobank. The remaining five recruited during the COVID-19 pandemic had no self-reported symptoms or positive PCR, four of them subsequently became infected, and their post-infection samples were included in the CP cohort. Detailed information about the donors is provided in Table S1.

We tested all individuals for the presence of IgGs to the receptor-binding domain (RBD) and T cell response to pools of peptides derived from the Membrane protein (M), Nucleoprotein (N), and Spike protein (S).

A humoral response was induced in the majority of individuals: at TP1, 88% of CDs had detectable levels of anti-RBD IgG antibodies (**Fig. 1A**). Individual antigens demonstrated comparable levels of T cell reactivity: 80–90% of CDs had detectable specific T cells depending on the tested proteins (**Fig. 1B**). However, overall T cell reactivity was more frequently observed than humoral response: only three (6%) individuals lacked T cell reactivity to any of the tested antigens versus six (12%) seronegative patients. Accordingly, in five out of six seronegative individuals, we detected T cell responses to two or three individual antigens. Of the three donors without T cell reactivity, a humoral response was detected in two (**Fig. 1C**).

**Figure 1.**
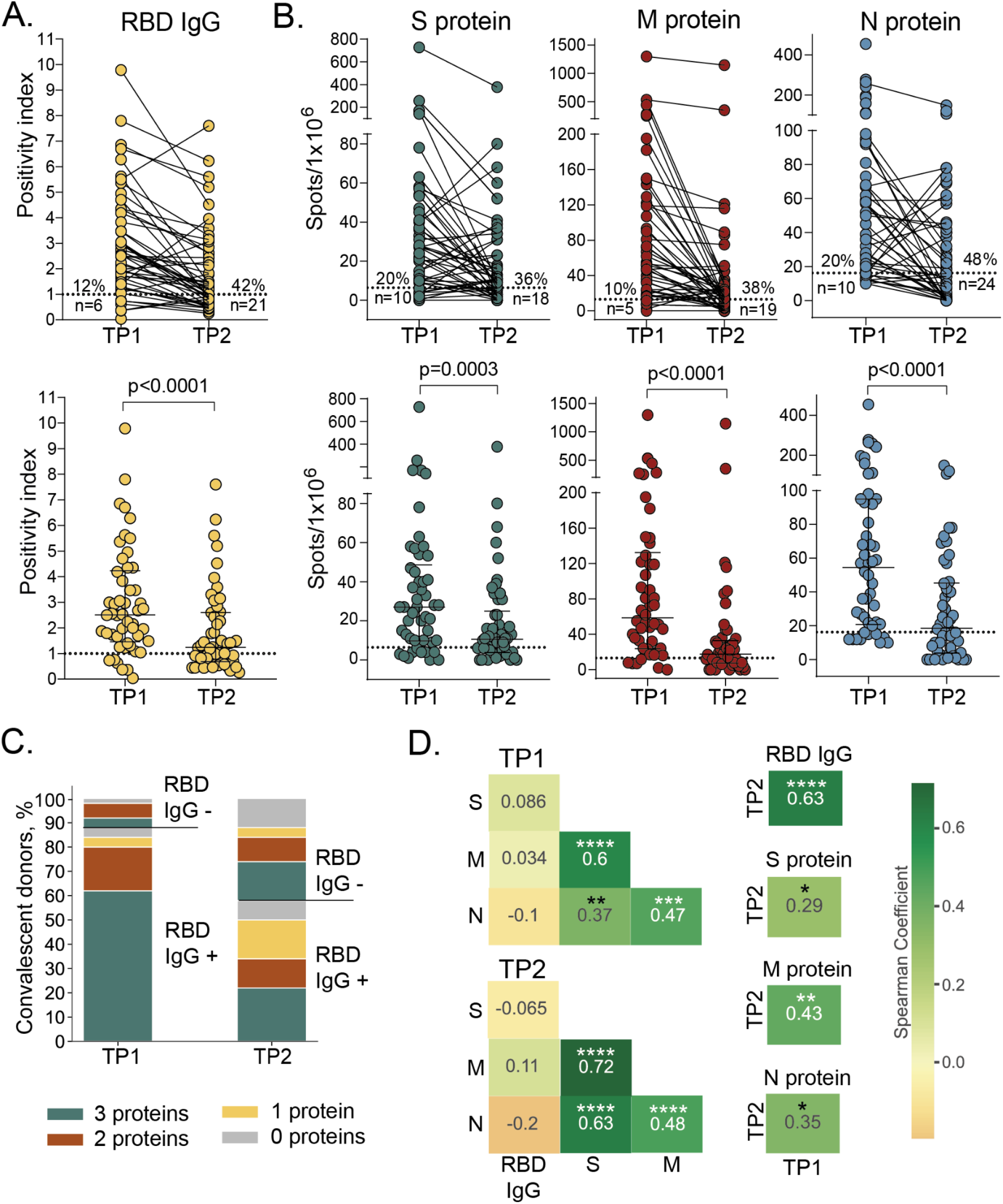
Persistence of humoral and cellular response in CP cohort. **A**. Levels of anti-RBD IgG as measured by ELISA at the two time-points. Means of two independent measurements are plotted. Upper plot shows samples from the same donor, connected by lines. Lower plot shows median with interquartile range (paired Wilcoxon test). **B**. Magnitude of T cell response to pools of peptides derived from S (green), M (red), and N proteins (blue) as measured by IFNγ ELISpot. Means of two independent measurements with negative control subtracted are plotted. Upper and lower plots are presented as in **A**. Dotted lines in **A** and **B** mark cut-offs, the number and share of individuals without detectable response is indicated. N = 50 for both **A** and **B. C**. Distribution of immune responses in CDs. Colors indicate T cell response to 0–3 peptide pools; RBD IgG+ and RBD IgG-respectively indicate presence and absence of anti-RBD IgG. **D**. Spearman correlation between humoral and cellular responses to different SARS-CoV-2 antigens. *p ≤ 0.05; **p ≤ 0.01; ***p ≤ 0.001; ****p ≤ 0.0001

After eight months, the immune response to SARS-CoV-2 decreased dramatically. Only 29 (58%) CDs maintained detectable IgG levels at the TP2 (**Fig. 1A**). T cell response also significantly diminished over time: depending on the tested antigen, specific T cells could not be detected at TP2 in 18–24 (36–48%) individuals, compared to 5–10 (10-20%) at TP1. N protein was characterized by the lowest number of responders at TP2 (**Fig. 1B**). Nevertheless, overall T cell reactivity to any of the antigens was more persistent than antibody response: 10 (20%) individuals lacked detectable T cells compared with 21 (42%) seronegative individuals at TP2. Notably, eight of the 21 seronegative CDs had measurable T cell responses to all three antigens (**Fig. 1C**). T cell responses to the S, M and N proteins were correlated regardless of the time-point. The response to each protein correlated between two time-points (**Fig. 1D, S1**). Antibody levels correlated between two time-points and did not correlate with T cell responses to all antigens. We did not observe any correlation between antibody titer and age or sex of donors or period of blood sampling within either TP1 or TP2 (**Fig. S2A, B**); similarly, the number of antigen-specific T cells was not significantly influenced by age or sex of donors or time since disease onset (**Fig. S2D-F**).

Some donors demonstrated an increase of response at TP2, including six in whom we observed an increase of T cell responses to two or three antigens (**Fig. S2G**). One of the explanations is the contact with SARS-CoV-2 between samplings.

### Epitope-specific CD8^+^ T cells response decreases but remains detectable over eight months

To study T cell response at the level of individual epitopes, we selected 20 CD8^+^ epitopes presented by common HLA I alleles and derived from different SARS-CoV-2 proteins, and which were previously described as immunogenic (**Table 1**). For 15 epitopes, we successfully folded peptide-MHC (pMHC) complexes. Based on the HLA genotype, we selected a subgroup of 26 donors (**Table S2**) and tested 4–15 donors (median = 8.5) for each epitope. To increase the frequency of epitope-specific memory T cells, we performed rapid *in vitro* antigen-specific memory cell expansion (Danilova et al., 2018; Shomuradova et al., 2020; Titov et al., 2021). Epitope-specific cells were further detected by flow cytometry using MHC tetramers (**Fig. S3**). Each expansion was performed in triplicate, and we used the number of positive wells as a surrogate indicator of T cell frequency.

**Table 1.**
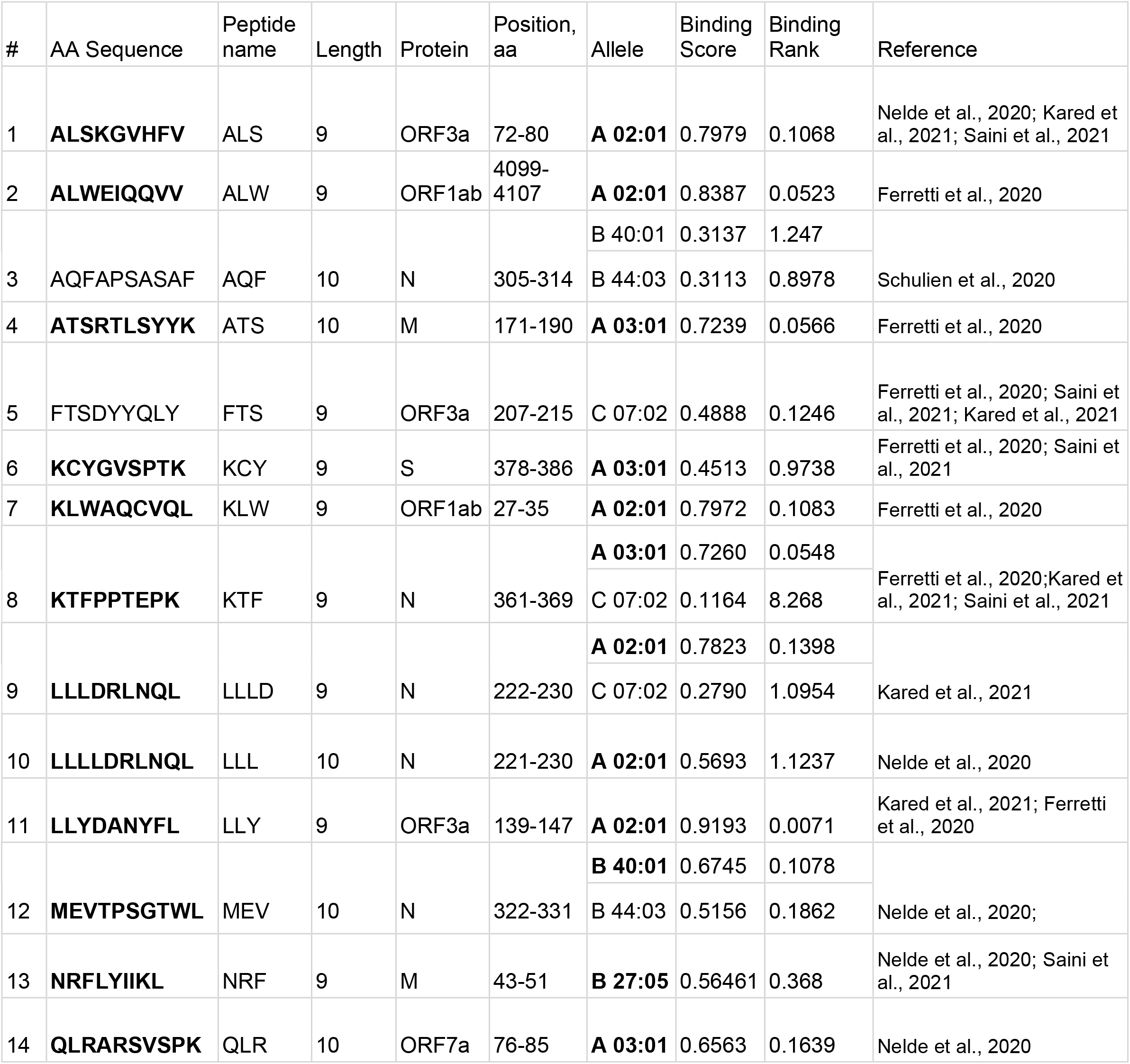
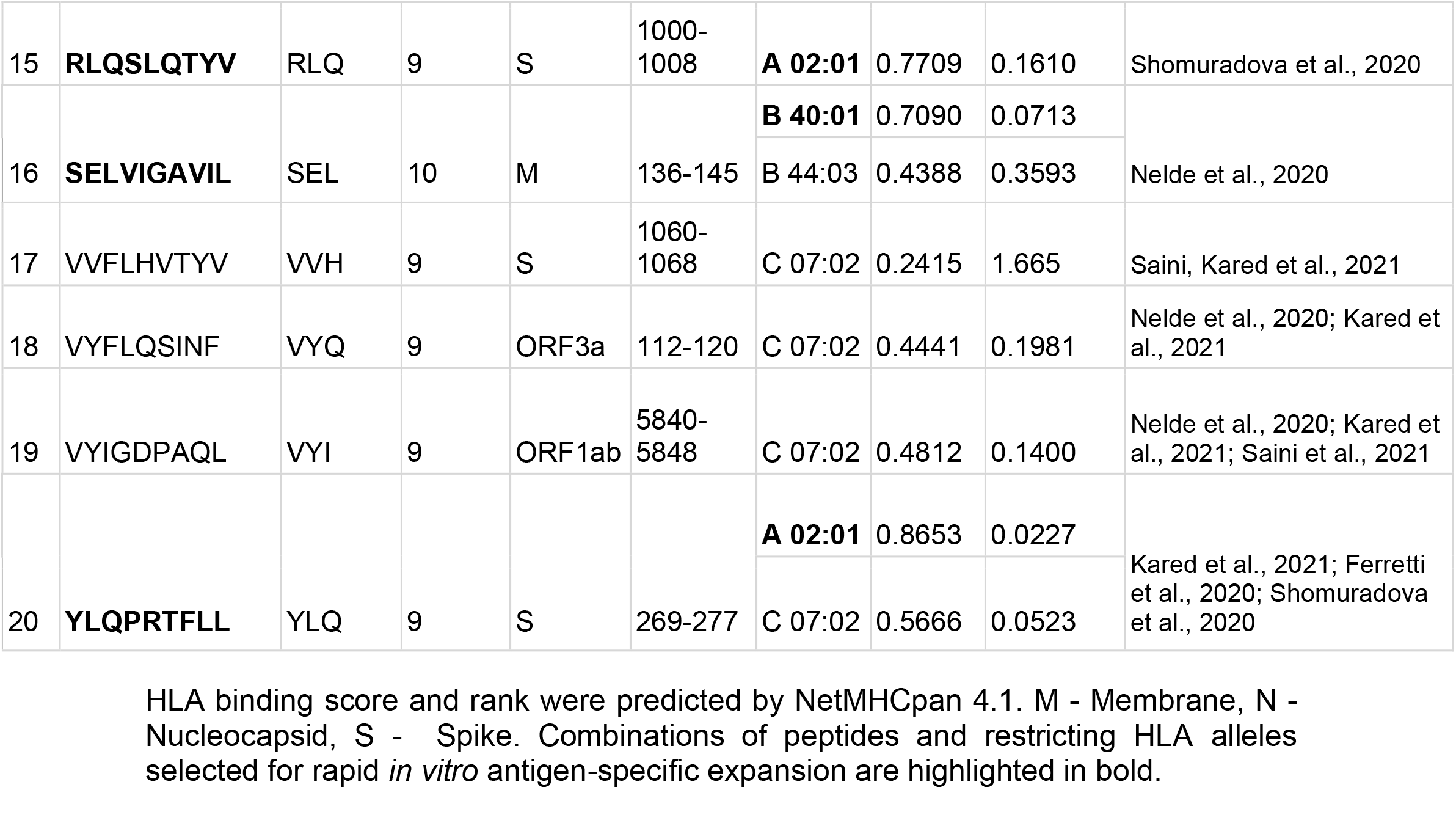
Peptides of SARS-CoV-2 used in this study.

We confirmed immunogenicity of twelve out of the 15 tested epitopes, and 11 of them were immunodominant, with epitope-specific cells detected in more than 50% of donors. Six epitopes (YLQ, ALW, LLY, KCY, KTF, and MEV) induced reactivity in 100% of CDs with the relevant HLA allele, and the response to four of them was detected in 3/3 wells (**Fig. 2A**). Only one epitope (LLY) yielded a weak response in a single healthy donor. At TP2, T cells specific to all 12 immunogenic epitopes were still detectable in at least a share of the patients, but the number of recognized epitopes per patient decreased significantly (**Fig. 2B**). Reduced frequency of specific T cells was characteristic of individual epitopes rather than patients: in most individuals, only a subset of the epitopes lost their immunogenicity (**Fig. 2C**; **Fig. S3**). In two donors (p1495 and p1426), we detected almost complete disappearance of antigen-specific cells, whereas in donor p1445, these cells were more abundant at TP2.

**Figure 2.**
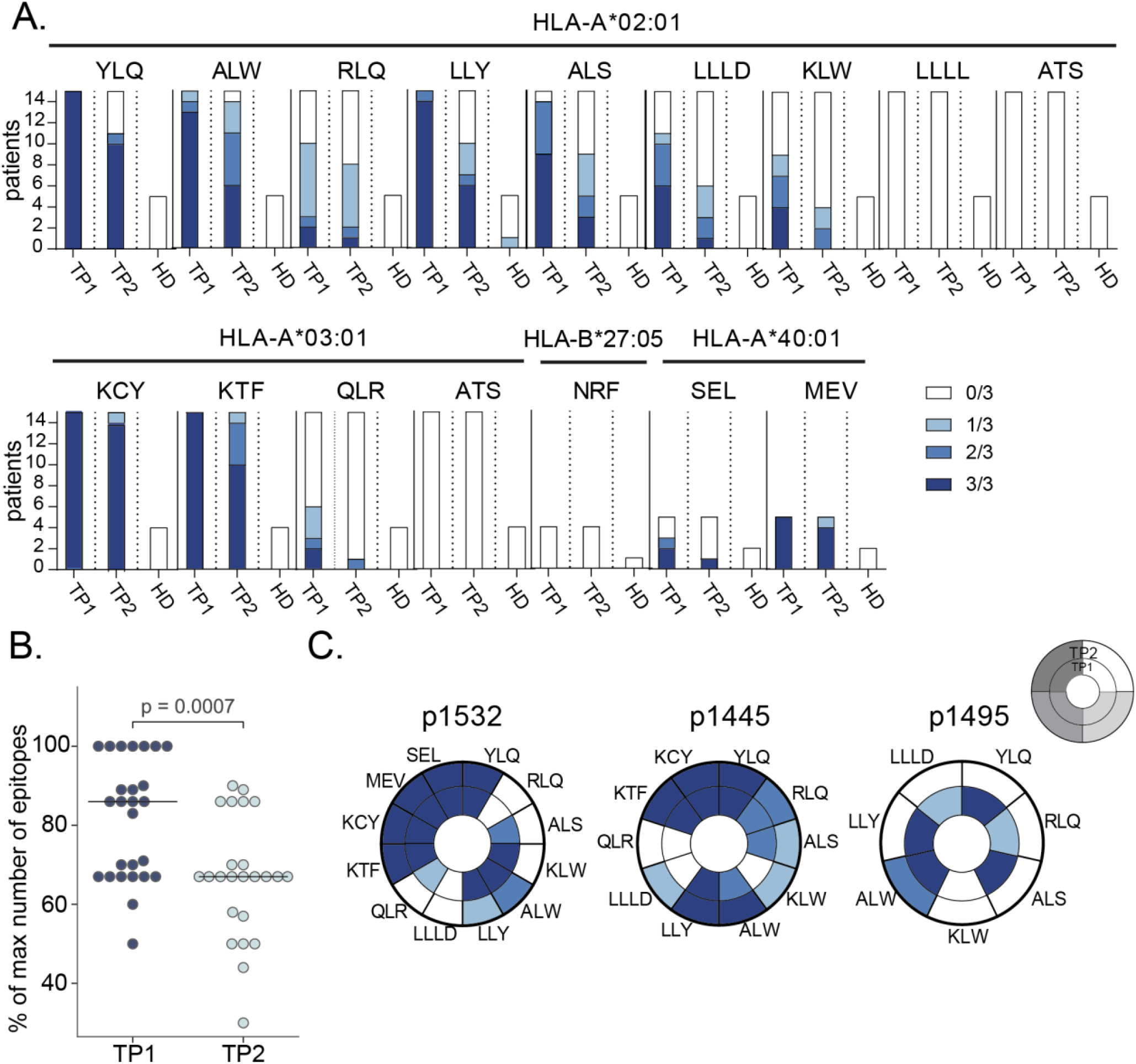
Frequency of epitope-specific T cells in CDs and healthy donors (HD). **A**. Number of wells with detected epitope-specific CD8^+^ cells after rapid *in vitro* expansion of PBMCs from CP (n = 26) and HD (n = 7). Bar height represents the number of HLA-allele carriers tested for response to each epitope. Color intensity indicates the number of wells containing MHC-tetramer^+^ cells. Epitopes represented by three or four-letter codes and their restricting HLA alleles are listed at top. **B**. Proportion of recognized epitopes out of the total number of tested epitopes—*i*.*e*., epitopes presented by the individual’s HLA alleles. Each dot represents one donor, response in at least one well is considered positive. Paired Mann-Whitney test, with p-value indicated above. **C**. Changes in the detection of antigen-specific cells after rapid *in vitro* expansion in TP1 (inner circle) versus TP2 (outer circle) for three representative donors (p1532, p1445, p1495). Each segment corresponds to epitopes indicated by three or four-letter codes.

### Dominant epitope-specific CD8^+^ T cell response arises from clonal diversity

We analyzed the repertoires of TCRβ chains of MHC-tetramer^+^ populations and total peripheral blood mononuclear cell (PBMC) fractions by high-throughput sequencing. Epitope-specific T cell clonotypes were defined as sequences that were strongly (≥10-fold) and significantly (p<10^−12^, Fisher’s exact test) enriched in the MHC-tetramer^+^ population (**Fig. 3A**). We identified 756 clonotypes specific to nine epitopes. Of all the epitopes, KCY and KTF had the most diverse response, with a median of 27 clonotypes per patient for both (**Fig. 3B**). The epitopes characterized by the highest clonality were also those for which we observed the most robust response (**Fig. 2A**). The number of T cell clonotypes specific for KCY and KTF significantly decreased in TP2 (**Fig. 3B**). We observed a similar reduction in diversity when we compared the response to all epitopes. The median number of clonotypes per epitope was 13 and 5 in TP1 and TP2, respectively (**Fig. 3C**). Nevertheless, for all epitopes except RLQ, some epitope-specific clonotypes were detectable at TP2. The initial disparity in the number of specific clonotypes per epitope became less prominent at TP2 (**Fig. 3B**). We did not find a correlation between the diversity of epitope-specific responses at the two time-points (**Fig. S5A**). The number of clonotypes decreased at TP2 even for the epitopes that produced a response in 3/3 wells (**Fig. S5B**). Nevertheless, diverse clonal structure was associated with better survival of epitope-specific T cell response; epitopes with more stable response (*i*.*e*., the same number of wells in which MHC-tetramer^+^ cells were detected at both TP1 and TP2) were characterized by a higher number of epitope-specific clones at TP1 (**Fig. 3D**). The described epitope-specific T cell clonotypes were either undetectable or were observed at a very low frequency in the total TCR repertoire at both time-points. Only a few clonotypes were present at a frequency of >10^−4^ of all clonotypes (**Fig. 3E**). Nevertheless, the KCY, KTF, YLQ, and ALW epitopes—for which dominant responses were retained at TP2 (Fig. 2A)—all had more frequent specific clonotypes detectable at TP1 (**Fig. 3E**).

**Figure 3.**
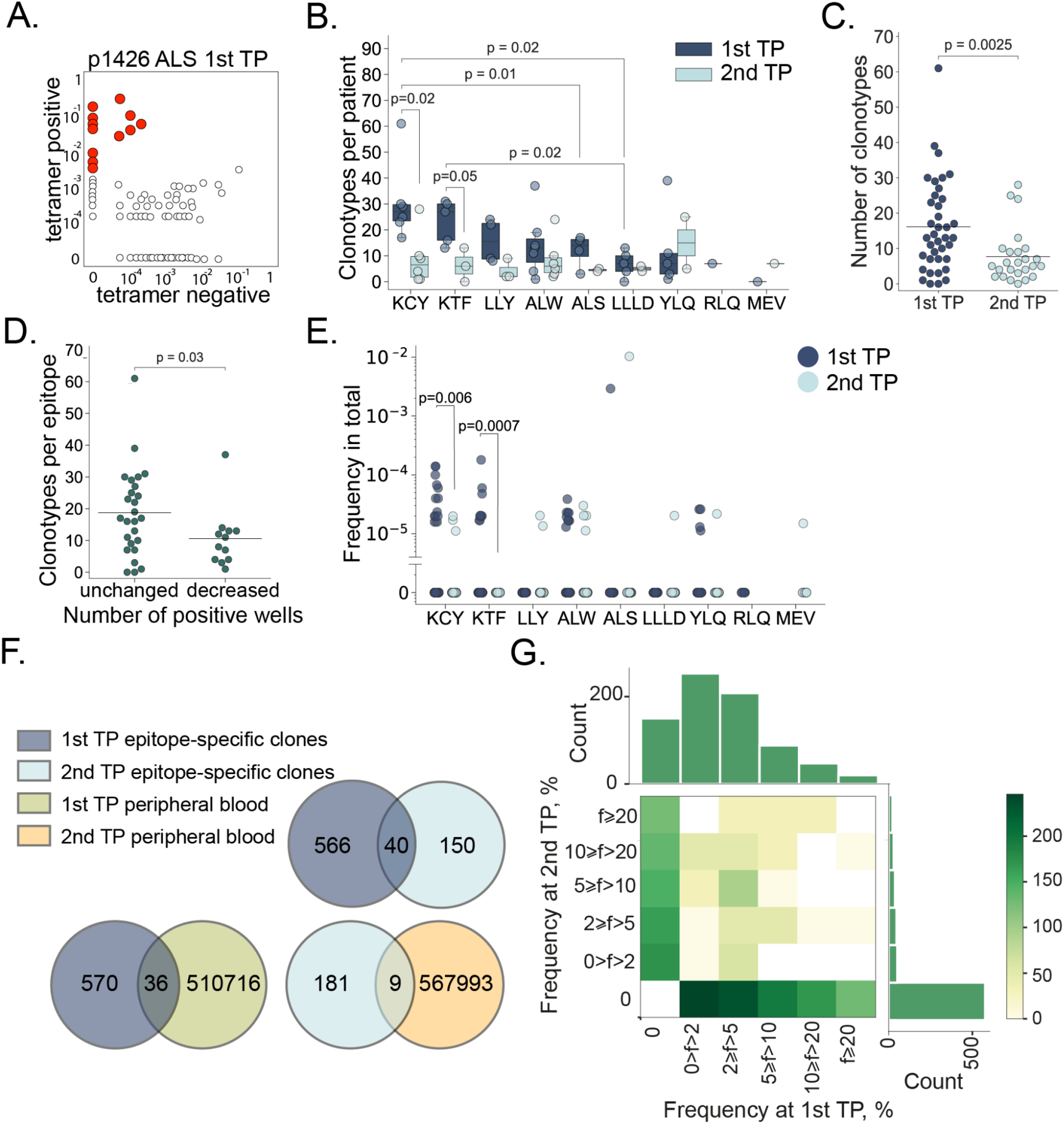
Clonal structure of the SARS-CoV-2 epitope-specific T cell response. **A**. MHC-tetramer^+^ CD8^+^ cells were subjected to fluorescence-activated cell sorting (FACS) after rapid *in vitro* peptide-induced expansion of PBMCs from CP (n = 14), and their TCRβ repertoire was sequenced. A representative enrichment plot is shown for patient p1426, epitope ALS at TP1, showing clonotype frequencies in MHC-tetramer^+^ CD8^+^ and MHC-tetramer^−^ CD8^+^ populations. Red dots represent clonotypes that are strongly (>10-fold) and significantly (p < 10^−12^, Fisher’s exact test) enriched in the MHC-tetramer^+^ population. **B**. Numbers of specific clonotypes for each epitope at TP1 (dark blue) and TP2 (light blue). Boxplots show the first and third quartiles with median; whiskers show 1.5 interquartile range values. **C**. Number of epitope-specific clonotypes in the CP cohort (n = 14), where each dot represents a unique patient-epitope combination (medians are shown by horizontal line). **D**. Number of specific clonotypes per epitope at TP1 which exhibited an unchanged or decreased overall response at TP2. **E**. Frequency of epitope-specific clonotypes in PBMC at TP1 (dark blue) and TP2 (light blue). **F**. Venn diagrams showing overlap between different clonotype fractions from the CP cohort. **G**. Frequency (f) of epitope-specific clonotypes in expansions performed at the two time-points. Color represents the number of clonotypes in each bin. Mann-Whitney p-values are shown only where significance was reached.

The clonal repertoire of epitope-specific CD8^+^ cells was markedly different between the two time-points. Of the 756 discovered epitope-specific clonotypes (including public clonotypes), only 40 (5.3%) were found at both time-points (**Fig. 3F**). This corresponded to 0–5 (median = 1.5) persistent clonotypes per patient. In order to rule out the possibility that this negligible overlap was the result of overly stringent criteria for designating clonotype specificity, we searched the epitope-specific clonotypes of TP1 in the total MHC-tetramer^+^ fraction of the TP2, and vice versa. This recovered 45 additional clonotypes, bringing the share of persistent clonotypes to 11.2% (**Fig. S5D**).

There was no obvious correlation between the relative clonotype abundance in the TP1 T cell expansion and its presence or abundance in the TP2 T cell expansion (**Fig. 3G**; **Fig. S6**). Likewise, we did not observe a correlation between clonotype persistence and the frequency with which it appeared in the repertoire at TP1 (**Fig. S6**). Only 5.9% and 4.7% of the epitope-specific clones from TP1 and TP2, respectively, were observed in the total PBMC repertoire, and we only observed 3% overlap in clonotypes from the total PBMC repertoires at the two timepoints (**Fig. 3F**; **Fig. S5C**).

### The persistence of epitope-specific T cells arises from response diversity

Out of the seven epitopes for which TCRs were obtained from more than one donor, LLY and YLQ demonstrated the highest similarity in their epitope-specific CDR3β as measured by the share of sequences belonging to clusters formed by the sequences with one or two amino acid substitutions (**Fig. 4A, B**). When only a single amino acid substitution or indel was permitted, 59.7% of the LLY-specific clonotypes were assigned to a cluster, versus 10.7% and 6.2% for KTF and KCY-specific clonotypes, respectively, which were characterized by the lowest similarity. Of the 715 unique epitope-specific clonotypes, 19 were public, with the same CDR3β amino acid sequence shared by 2–4 individuals in our cohort (some were also encoded by multiple nucleotide sequences in one individual). Epitope LLY was recognized by the greatest number of public clonotypes—eight, corresponding to 43.1% of all LLY-specific clonotypes—while KTF and KCY had only one and two, respectively (**Fig. 4C**).

**Figure 4.**
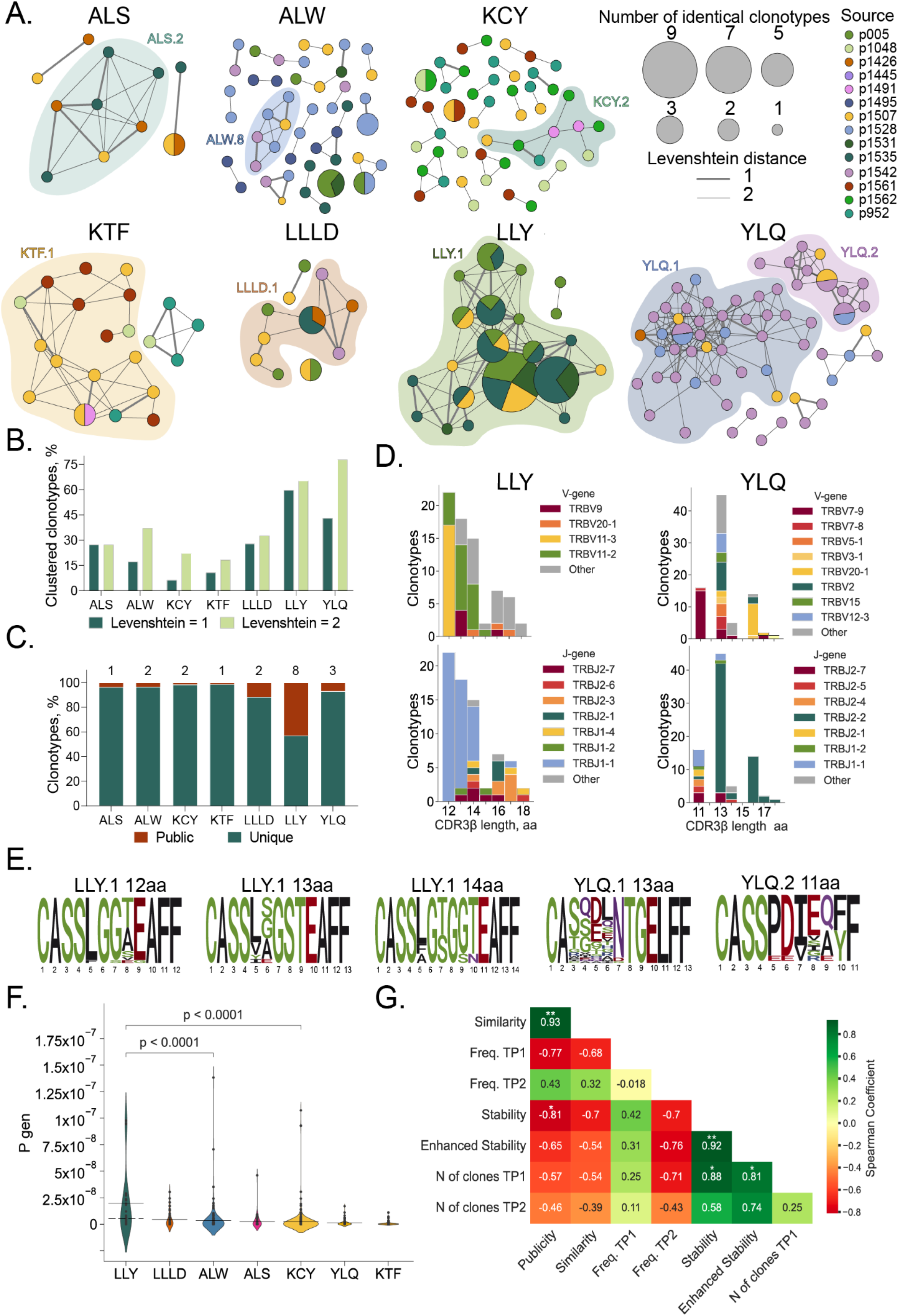
Clonal structure of SARS-CoV-2 epitope-specific response. **A**. Clusters of CDR3β sequences of epitope-specific CD8^+^ T cells. Each node represents a unique CDR3β amino acid sequence or public sequences (CDR3β amino acid sequences shared by 2–4 individuals in our cohort), where the node size is proportional to the number of identical clonotypes. Lines connect similar CDR3β sequences, where the thickness indicates Levenshtein distance = 1 or 2. Colors indicate donors. Only clusters with two or more members are shown. Clusters with 6 or more nods highlighted with color. **B**. Proportion of clustered epitope-specific T cell clonotypes. **C**. Fraction of public (dark red) and unique (dark green) epitope-specific clonotypes. Numbers of public clones are listed at top. **D**. Histograms of V (upper plots) and J gene usage (lower plots) usage in LLY-(left) and YLQ-specific clones (right). **E**. Position-weight matrices for CDR3β sequences with the most common length observed in LLY and YLQ clusters. Cluster numbers corresponds to numbers shown in **A. F**. Probability of V(D)J-recombination with the observed V- and/or J-gene usage as calculated by the OLGA algorithm (Sethna et al., 2019). Solid line represents the mean and dashed line represents the median Pgen. Mann-Whitney test, p-values are indicated. **G**. Spearman correlation between various parameters of SARS-CoV-2 epitope-specific T cell response. *p ≤ 0.05, **p ≤ 0.01, ***p ≤ 0.001, ****p ≤ 0.0001.

Next, we compared CDR3β regions with specificity for LLY, YLQ, KTF, LLLD and ALS epitopes with CDR3β sequences annotated as recognizing the same epitopes in the Multiplex Identification of T cell Receptor Antigen Specificity (MIRA) dataset (Snyder et al., 2020) and the VDJdb database (http://vdjdb.cdr3.net) (Shugay et al., 2017; Bagaev et al., 2019). Both datasets contain CDR3β sequences that are highly similar to the sequences obtained in this study (**Fig. S7A**). Neither MIRA nor VDJdb contained TCRs annotated as being specific for ALW or KCY epitopes. Nevertheless, we found some similar (max Levenshtein distance = 1) CDR3β sequences that were annotated as being specific for other SARS-CoV-2 epitopes (**Fig. S7A**). A similarity between CDR3β regions recognizing different epitopes could be explained by the impact of TCRɑ pairing or germline-based recognition mediated by CDR1 and CDR2. We found that 32% of the ALW-specific clonotypes described in this study were similar to YLQ-specific CDR3β sequences from the MIRA database (includes sequences specific to <u>YLQ</u>PRTFL, <u>YLQ</u>PRTFLL, and YYVG<u>YLQ</u>PRTF peptides). High promiscuity was also observed for KCY, where 83 (26.9%) epitope-specific clonotypes had CDR3β sequences similar to TCRs with another specificity. Most YLQ- and KTF-specific CDR3β sequences clustered with CDR3β annotated with the same specificity in the other databases (**Fig.S7A, B**).

It was previously shown that epitopes of 9–10 amino acids (AA) are mostly recognized by CDR3β regions with a length of 13–15 AA. This CDR3β length is associated with public TCRs with the highest V(D)J-rearrangement probability (Pogorelyy et al., 2018). The clonotypes specific to most of the SARS-CoV-2 epitopes obtained in this study followed this trend, with the exception of ALW and LLY. ALW-specific clonotypes had the longest CDR3β, with 16 AA being the most common length; in contrast, the majority of LLY-specific clonotypes had CDR3β of 12 AA. We did not observe YLQ-specific clones with lengths of 12 and 15 AA (**Fig. 4D, Fig. S8A**) because of their rarity. These lengths were observed in just 2.2% and 5%, respectively, of the 821 YLQ-specific CDR3β regions annotated in VDJdb. LLY-specific clonotypes with a short (12–14 AA) CDR3β most often used TRBV11-3 and TRBV11-2, while YLQ-clonotypes with 13 AA CDR3β were formed by a large variety of TRBVs. Clonotypes specific to both LLY and YLQ were highly biased in J-gene usage, with a predominance of TRBJ1-1 and TRBJ2-2, respectively (**Fig. 4D, Fig. S8A**). The high diversity of V gene usage by YLQ-specific clones is explained by the substantial role of TCRɑ in epitope recognition, as we demonstrated previously (Wu et al., 2021).

Position-weight matrices of the CDR3β regions with the most common lengths demonstrated more diversity in YLQ-specific than LLY-specific TCRβ repertoires (**Fig. 4E**). YLQ-specific clonotypes with a length of 11 (from cluster YLQ.2) or 13 AA (from cluster YLQ.1) had the same CDR3 motifs as those we described earlier (Shomuradova et al., 2020). LLY-specific clones with a length of 14 AA contained the CDR3 motif CASS[LFA]G[TS]G[GS][TN]EAFF described in (Francis et al., 2021), while clones with lengths of 12 and 13 AA had previously undescribed CASS[LF]G[GS][TASVE][EG]AFF and CASS[LVYI][SGAE]GSTEAFF CDR3β motifs, respectively. Moreover, we identified CDR3β motifs recognizing ALS, ALW, KCY, KTF, and LLLD (**Fig. S8B**). The high similarity of the LLY-specific clonotypes and abundance of public CDR3β sequences is a result of the high probability of V(D)J-recombination (Pgen). The mean Pgen as calculated by the OLGA algorithm (Sethna et al., 2019) restriction was about 1.97 × 10^−8^ (max Pgen = 9.77 × 10^−8^) (**Fig. 4F**).

In order to elucidate the factors contributing to the longevity of response to a particular epitope, we measured the correlation of publicity (share of clones found more than one donor); similarity (share of clones belonging to a cluster); average frequency of epitope-specific clonotypes in the total repertoire of TP1 or TP2; average number of clones at TP1; and finally, stability and enhanced stability, which are respectively defined as the proportion of non-zero and 3/3 epitope-specific responses at TP2. The quantity of epitope-specific clones at TP1 strongly and significantly influenced both stability and enhanced stability (**Fig. 4G**). This highlights the importance of induction of polyclonal response to the formation of long-term T cell memory.

## Discussion

We have studied the durability of humoral and cellular immune responses to SARS-CoV-2 by analyzing paired blood samples of 50 convalescent patients collected at 17–72 days and 180–292 days post-infection. Early on, T cell response was more commonly observed than humoral response, and most seronegative individuals had measurable T cell responses to multiple antigens. In combination with data from other studies (Nelde et al., 2020; da Silva Antunes et al., 2021; Oberhardt et al., 2021), this indicates that T cell response serves as a more reliable measure of past COVID-19 infection. After a median follow-up time of 8 months, we observed a decrease in the SARS-CoV-2-specific immune response. This is in line with other studies, where the average half-life time of antibody and T cell response was 6–8 months and 10–15 months, respectively (Dan et al., 2021; Sherina et al., 2021; Jung et al., 2021, Marcote et al., 2021). We also confirmed that the humoral response is less durable and diminishes faster. Cellular immunity against SARS-CoV-2 was present in the great majority of adults at 6 months following mild and moderate infection, as was also shown by (Jung et al., 2021). Of the three structural antigens examined here, N protein was the least immunogenic, as shown previously (Shomuradova et al., 2020; Thieme et al., 2020; Titov et al., 2021), and the response to it was the least stable. Anti-RBD antibody levels did not correlate with T cell response in this study, which might be characteristic of the RBD antigen. In contrast, other studies report a strong correlation of humoral response with T cell frequencies (Chen et al., 2021; Molodtsov et al., 2021).

The focus of this work was the analysis of CD8^+^ T cell response to individual epitopes at the clonotype level. We tested the immunogenicity of 15 known epitopes in 26 donors. In order to confirm their immunogenicity, detect minor T cell clones, and describe the structure of the antigen-specific repertoire, we used a highly sensitive method—rapid *ex vivo* T cell expansion. We divided the cells equally between three wells and used the number of wells containing MHC-tetramer^+^ T cells after expansion as a surrogate marker of T cell frequency. In our study, the previously reported epitopes LLLLD, ATS and NRF were not immunogenic. The remaining epitopes differed in their immunogenicity; 11 were immunodominant with the response observed in more than 50% of CDs. Only one epitope (LLY) yielded a response in a healthy donor, which is in agreement with the other studyreporting that epitope as cross-reactive (Francis et al., 2021). At 180 to 292 days post-infection, epitope-specific T cells were still detectable, although the number of recognized epitopes decreased as well as the number of detectable epitope-specific clonotypes and their frequency. Importantly, the change in response magnitude was not universal across epitopes, with some retaining high immunogenicity for 8 months. Thus, antigen-specific T cells not only formed long-living memory populations in blood, but also proliferated after stimulation with their antigen. This finding corresponds to previous work showing the detection of SARS-CoV-1 memory T cells after 17 years (Le Bert et al., 2020) and the presence of SARS-CoV-2-specific memory B and T cells in the majority of a cohort of patients followed up at 6–15 months post-infection, with a reduction of the T cell response at 12–15 months (Marcotte et al., 2022). Our data confirmed that more abundant clonotypes do not generally persist better, and that clonal contraction and disappearance are more associated with a strongly proliferative phenotype (Adamo et al., 2021).

We described 715 unique clonotypes. Some epitopes had high clonal diversity (*e*.*g*., KCY had 223 unique clonotypes, and KTF and 130) while others were recognized by few clonotypes. Nevertheless, a decrease in the number of clones was characteristic even for polyclonal responses. Moreover, reduction in the number of clonotypes was shown even for high-frequency epitopes, although the clonality of the response correlated with the persistence of cells in the blood. It is worth noting that KTF and KCY were also characterized by the greatest number of specific clonotypes per patient and the most durable response. We can thus conclude that a high-diversity response contributes to persistence more than the abundance of the clones. We found very little overlap in terms of clonal representation between the two time-points. This is probably because T cell repertoires are highly dynamic and undergo major changes over the course of 6–9 months, and also because SARS-CoV-2-specific cells occupy a very small proportion of the repertoire and the probability of harvesting the same clones is low. The structure of the epitope-specific response also differed between epitopes. LLY was recognized by TCRs with the most mutual similarity, which accounted for 8 out of 19 public CDR3β sequences, while the rest of the epitopes had low levels of similarity of specific TCRs. Our work thus shows that the formation of a long-lasting immune response to SARS-CoV-2 is dependent on the clonal diversity of the initial response rather than the abundance of a particular clonotype in the peripheral circulation. The immunodominant T cell epitopes recognized by diverse T cell populations should be considered for inclusion in the new generation of vaccines.

## Resource availability

### Lead contact

Further information and requests for resources and reagent should be directed to and will be fulfilled by the lead contact.

### Materials availability

This study did not generate new unique reagents.

## Materials and methods

### Human subjects

50 COVID-19 convalescent donors (CD) from Moscow, Russia volunteered for participation in this study. COVID-19 was confirmed by positive SARS-CoV-2 RT-PCR test. All donors signed the informed consent form approved by the National Research Center for Hematology ethical committee (N 150, 02.07.2020) before enrollment. The severity of disease was defined according to the classification scheme used by the US National Institutes of Health (from www.covid19treatmentguidelines.nih.gov): asymptomatic (lack of symptoms), mild severity (fever, cough, muscle pain, but without respiratory difficulty or abnormal chest imaging) and moderate/severe (lower respiratory disease at CT scan or clinical assessment, oxygen saturation (SaO2) > 93% on room air, but lung infiltrates less than 50%). Additionally, 19 healthy donor (HD) samples were obtained: 14 from the National Medical Research Center for Hematology blood bank (cryopreserved no later than August 2019) with the approval of the local ethical committee, and 5 recruited during the COVID-19 pandemic with no self-reported symptoms and negative PCR test results (four of them subsequently became infected, and their post-infection samples were included in CP cohort).

### Peripheral blood mononuclear cell (PBMC) isolation

30 mL of venous blood from donors was collected into EDTA tubes (Sarstedt) and subjected to Ficoll (Paneco) density gradient centrifugation (400 x g, 30 min). Isolated PBMCs were washed with PBS containing 2 mM EDTA and used for assays or frozen in fetal bovine serum containing 7% DMSO.

### HLA genotyping

For most donors, HLA genotyping was performed with the One Lambda ALLType kit (Thermo Fisher Scientific), which uses multiplex PCR to amplify full HLAA/B/C gene sequences, and from exon 2 to the 3’^’^ UTR of the HLA-DRB1/3/4/5/DQB1 genes. Prepared libraries were run on an Illumina MiSeq sequencer using a standard flow-cell with 2 × 150 paired-end sequencing. Reads were analyzed using the One Lambda HLA TypeStream Visual Software (TSV), version 2.0.0.27232, and the IPD-IMGT/HLA database 3.39.0.0. Other donors were HLA genotyped by Sanger sequencing for loci HLA-A, B, C, DRB1, and DQB1 using Protrans S4 and S3 reagents. PCR products were prepared for sequencing with BigDye Terminator v1.1 (Thermo Fisher Scientific). Capillary electrophoresis was performed on a Nanophore 05 Genetic Analyzer.

### SARS-CoV-2 peptides

Putative 20 epitopes of SARS-CoV-2 proteins were included in the analysis if they were binders (rank < 2) according to NetMHCpan 4.1 (Reynisson et al., 2020) and immunogenic according to published data. Detailed information about the selected peptides is listed in **Table 1**. Peptides (at least 95% purity) were synthesized either by Peptide 2.0 or by the Shemyakin-Ovchinnikov Institute of Bioorganic Chemistry RAS. Peptides containing Cys and/or Met were diluted in a PBS/isopropanol mixture (1:1 v/v) at concentrations of up to 10–25 mM. Other peptides were diluted in DMSO (Sigma-Aldrich) at up to 30–40 mM.

### Antigen-specific T cell expansion

For rapid *in vitro* expansion, we used PBMCs of donors with 1–4 of the HLA-A*02:01, HLA-A*03:01, HLA-B*40:01, and HLA-B*27:05 alleles, as previously described (Danilova et al., 2018). Briefly, 9 × 10^6^ cells were split between three wells and incubated for 10–12 days in RPMI 1640 culture medium supplemented with 10% normal human A/B serum, 1 mM sodium pyruvate, 25 ng/mL IL-7, 40 ng/mL IL-15, and 50 ng/mL IL-2 (Miltenyi) at a final volume of 2 ml/well. Half of the medium was replaced on days 3, 5, and 7. A mix of patient HLA-specific peptides (**Table 1**) of SARS-CoV-2 in DMSO or MES buffer / isopropanol mixture was added on day 0. The final concentration of each peptide in the medium was 10 ng/mL.

### Production of biotinylated MHC class I/peptide complexes

All chemicals used were of analytical grade and were purchased from Sigma-Aldrich except for EDTA, sodium azide (Amresco), sodium chloride (Malinovoe Ozero) and protease inhibitor cocktail (Thermo). Recombinant biotin ligase used was homemade. Solutions were prepared on deionized water (Simplicity Water purification system, Merck-Millipore) and filtered using 0.45 um syringe filters (Sarstedt) or bottle top filters (Nalgene Nunc International).

Soluble HLA-A2 loaded with different peptides was prepared by *in vitro* folding of *E. coli* inclusion bodies as described previously (Garboczi et al., 1992) with modifications (Shomuradova et al., 2020). Briefly, human heavy (HLA-A*02:01 with biotinylation tag) and light (beta-2 microglobulin) chains were expressed in *E. coli* strain BL21(DE3) pLysS as inclusion bodies and used for *in vitro* folding. Peptides and light and heavy chains were mixed in folding buffer (100 mM Tris-HCl, 400 mM arginine, 5 mM reduced glutathione, 0.5 mM oxidized glutathione, 2 mM EDTA, protease inhibitors, 1 mM PMSF, pH = 8.0) at a 30:4:3 final molar ratio. Folding reactions were incubated at 8 °C for up to 5 days. Correctly-folded complexes were purified on a Superdex 75 pg 16/600 column (Cytiva) using Tris-buffered saline (20 mM Tris-HCl, 150 mM NaCl, pH 8.0) as mobile phase. Complexes were biotinylated by in-house-made biotin ligase (20 mM Tris-HCl, 150 mM NaCl, 40 mM ATP, 0.4 mM biotin, 6.5 mM MgCl2, 25 μg/ml biotin ligase, protease inhibitor cocktail) at 30 °C for 1 hour or at 8 °C overnight and purified on a Superdex 75 pg 10/300 column. Biotinylated monomers were concentrated to a final concentration of 0.4–1.0 mg/ml and stored in 20% glycerol, 0.1% sodium azide, 0.1 mM EDTA, and protease inhibitor cocktail. Concentrations were determined using specific absorbance, with A280 = 2.36 and 1.68 for HLA-A and hB2M, respectively (as calculated in SnapGene Viewer based on amino acid sequence). Plasmids encoding HLA and b2-microglobulin were kindly provided by Ton Schumacher (The Netherlands Cancer Institute, Amsterdam, Netherlands).

### MHC-tetramer staining

Antigen-specific cells were detected by staining with CD3-AF700, CD8-FITC, 7AAD, and with combinations of two different peptide-MHC-tetramer complexes conjugated with streptavidin-allophycocyanin and streptavidin-R-phycoerythrin (Thermo Fisher Scientific) as previously described (Vdovin et al., 2016). We considered a sample well as positive if the percent of MHC-tetramer^+^ CD3^+^CD8^+^ cells was greater than 0.03– 0.4%, depending on the MHC-tetramer. An Aria III cell sorter (BD Biosciences) was used to sort cells, and data were analyzed using FlowJo Software (version 10.6.1) .

### ELISA

We used the IVD ELISA kit (National Research Center for Hematology) for the detection of anti-RBD IgG according to the manufacturers’ instructions. The optical density (OD) was measured at 450 nm with a reference of 650 nm on a MultiScan FC (Thermo Fisher Scientific) instrument. The mean of two OD values for each sample was divided by the mean of two OD values of a positive control and used as a positivity index. The cut-off was determined according to manufacturers’ instructions, and all samples with positivity index >1 were considered positive.

### IFNγ ELISPOT

IFNγ production by antigen-specific T cells was measured with the ImmunoSpot human IFNγ single-color ELISPOT kit (CTL) with a 96-well nitrocellulose plate precoated with the human IFNγ capture antibody. Thawed PBMCs were plated at a density of 5 × 10^5^ cells/well and pulsed with SARS-CoV-2 S, M or N protein-derived peptide pools (130-126-701, 130-126-703, 130-126-699, Miltenyi Biotec) separately in duplicates at a final concentration of 1 μM in serum-free testing medium (CTL) containing 1 mM GlutaMAX (GIBCO) at a final volume 200 μL/well. Plates were incubated for 16 h at 37 °C in 5% CO_2_. Assays were performed according to the manufacturer’s instructions. Spots were counted by CTL ImmunoSpot Analyzer using ImmunoSpot software. We subtracted the background (negative control) from each value and used the average value of two wells with the same peptide pool. We measured the T cell response of 11 HD to determine a cut-off for each peptide pool as mean + 1.69 SD.

### TCR repertoire sequencing

TCR libraries of MHC-tetramer^+^ and MHC-tetramer^-^ fractions were prepared as described previously (Zvyagin et al., 2017). Briefly, the cDNA synthesis reaction for TCR β chains was carried out with a primer to the C-terminal region and SMART-Mk, providing a 5′ template-switch effect and containing a sample barcode for contamination control as well as a unique molecular identifier. TCR libraries of total PBMC samples were generated using the human multiplex TCR Kit (MiLaboratories) according to the manufacturer’s instructions. Sequencing was performed with an Illumina MiSeq or NextSeq platform. TCR repertoire data were analyzed using MIXCR, MIGEC, and VDJtools software with default settings.

### TCR repertoire analysis

TCRs specific to SARS-CoV-2 epitopes were defined as clonotypes with frequencies significantly enriched in the MHC-tetramer^+^ fraction relative to the MHC-tetramer^−^ fraction (≥10-fold higher frequency, p-value <10^−12^, exact Fisher test). Clones with multi-epitope specificity were removed from the analysis. Epitope-specific TCR sequences were matched against VDJdb and ImmunoCODE datasets using the VDJmatch tool, with a maximum Levenshtein distance of 1. Graphs were plotted using “igraph” R package version 1.2.6. TCR logos were plotted using “ggseqlogo” package version 0.1.

### Quantification and Statistical Analysis

All data comparisons were performed using GraphPad Prism 8 software and python3. Peptides’ HLA binding affinity score and rank were predicted by NetMHCpan 4.1.

## Data Availability

All data produced in the present work are contained in the manuscript, except list of CDR3 sequences, which is available upon request.

## Acknowledgments

We would like to express our gratitude to all donors who volunteered for our study; to the nurses who performed the venipuncture (Yulia Fadeeva, Anastasya Nisanova, Valentyna Mirponova, and Lubov Piskunova); and to our colleagues Naina Shakirova and Ekaterina Khamaganova for their kind help with the experiments. We thank Michael Eisenstein for careful and thoughtful manuscript editing. Wet lab experiments and bioinformatic work (Figures 1–4, and S1–S7) were supported by the Russian Science Foundation grant 20-15-00395 (G.A.E.). I.Z. was funded by the Ministry of Science and Higher Education of the Russian Federation grant No. 075-15-2019-1789.

## Author contributions

Conceptualization, G.A.E., and K.V.Z.; Methodology, G.A.E., K.V.Z., I.Z.; Data Analysis, K.V.Z. and A.K.; Investigation, K.V.Z., A.K., S.A.S. and A.T.; Resources, K.V.Z., S.A.S., D.K. and O.V.S; Writing – Original Draft, K.V.Z.; Writing – Review & Editing, G.A.E., K.V.Z., A.K., S.A.S, A.T., I.Z.; Project Administration, G.A.E.; Funding Acquisition, G.A.E.

## Notes

### Competing Interest Statement

The authors have declared no competing interest.

### Funding Statement

The work was supported by the Russian Science Foundation grant 20-15-00395 (G.A.E.). I.Z. was funded by the Ministry of Science and Higher Education of the Russian Federation grant No. 075-15-2019-1789.

### Author Declarations

Ethics committee of the National research center for Hematology gave ethical approval for this work

