## Supplemental Files (Fig.S1-S8; Tab.S1-S2) for "Clonal diversity determines persistence of SARS-CoV-2 epitope-specific T cell response"

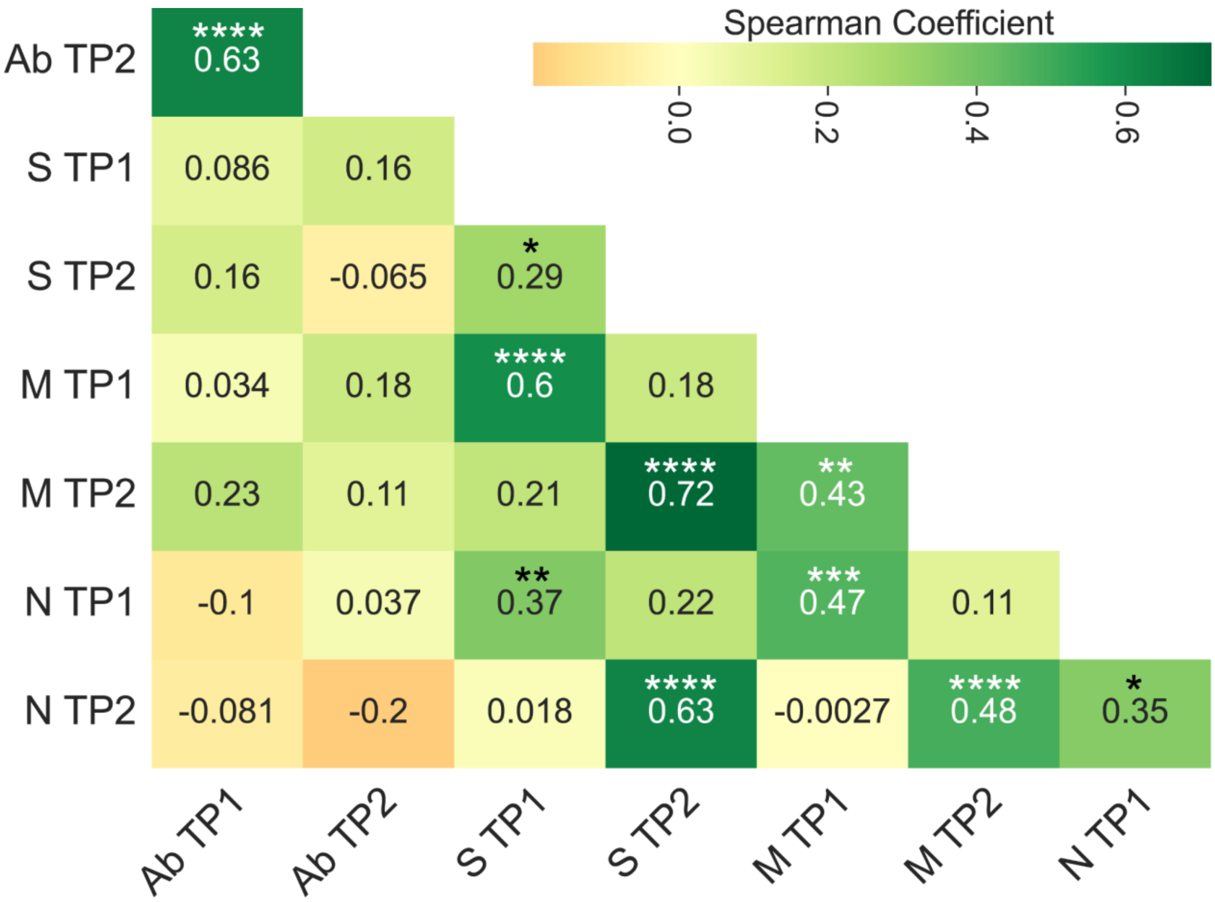


**Figure S1. Correlation of different parts of immune response.**
Spearman correlation between humoral (Ab) and cellular responses to different SARS-CoV-2 antigens (S, M, and N proteins).
*p ≤ 0.05; **p ≤ 0.01; ***p ≤ 0.001; ****p ≤ 0.0001

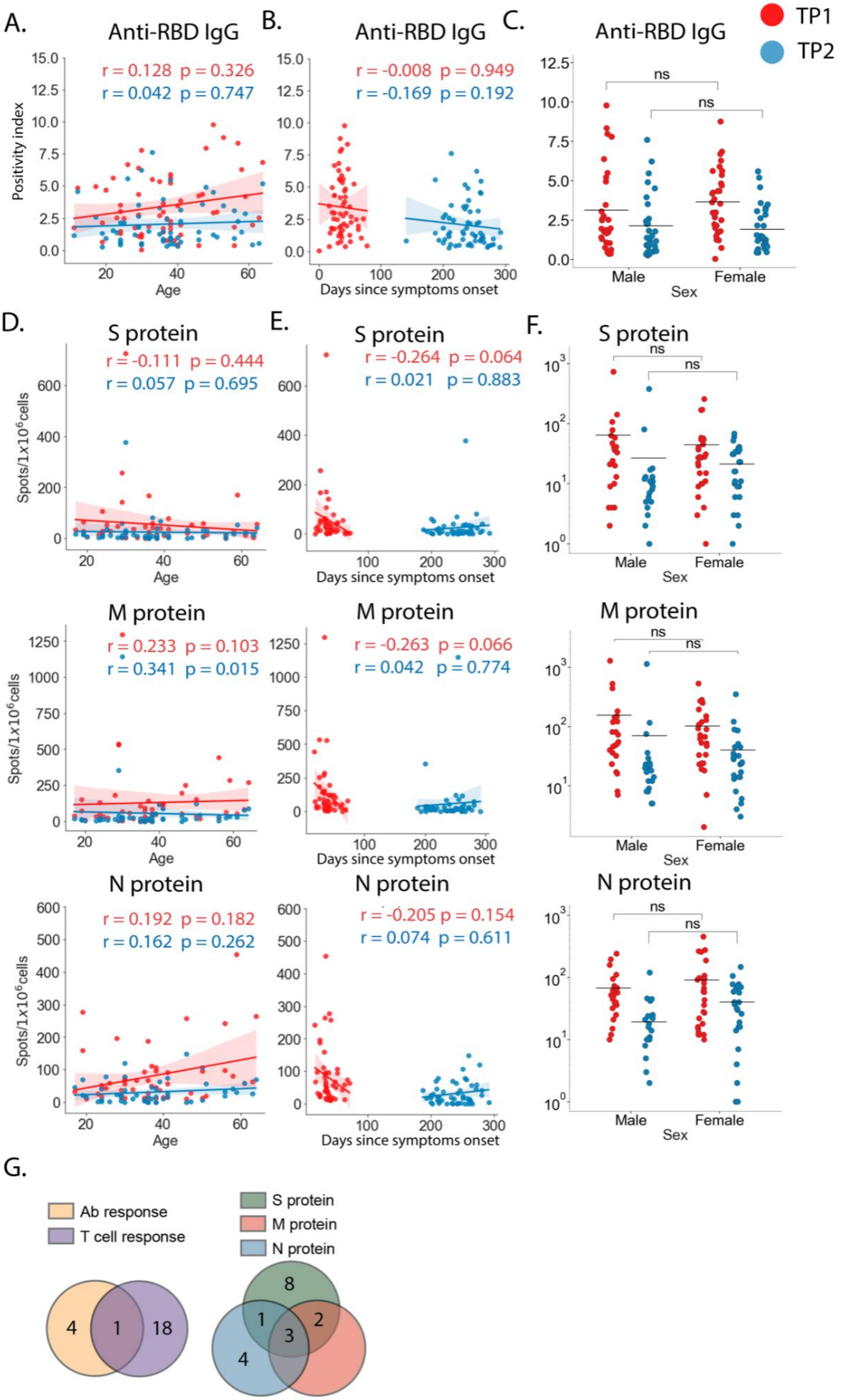
A.
Anti-RBD IgG
r = 0.128 p = 0.326
r = 0.042 p = 0.747
Positivity index
Age
B.
Anti-RBD IgG
r = -0.008 p = 0.949
r = -0.169 p = 0.192
Days since symptoms onset
C.
Anti-RBD IgG
ns
Male
Female
Sex
TP1
TP2
D.
S protein
r = -0.111 p = 0.444
r = 0.057 p = 0.695
Spots/1x10^6cells
Age
E.
S protein
r = -0.264 p = 0.064
r = 0.021 p = 0.883
Days since symptoms onset
F.
S protein
ns
Male
Female
Sex
M protein
r = 0.233 p = 0.103
r = 0.341 p = 0.015
M protein
r = -0.263 p = 0.066
r = 0.042 p = 0.774
M protein
N protein
r = 0.192 p = 0.182
r = 0.162 p = 0.262
N protein
r = -0.205 p = 0.154
r = 0.074 p = 0.611
N protein
G.
Ab response
T cell response
S protein
M protein
N protein
4
1
18
8
1
2
3
4

**Figure S2. Impact of donor age, sex, and sampling time-point on antibody and T cell response to different SARS-CoV-2 antigens. A.** Spearman correlation between donor age and antibody titers was measured by ELISA. **B.** Spearman correlation between time after the onset of the disease or positive PCR test and antibody titers. **C**. Effect of sex on antibody levels. **D.** Spearman correlation between donor age and magnitude of T cell response to S, M, or N protein as measured by IFN-γ ELISpot. **E.** Spearman correlation between time after onset of the disease or positive PCR test and magnitude of T cell response to S, M, or N protein. **F.** Effect of sex on the magnitude of T cell response to S, M, or N protein. r = correlation coefficient; Mann-Whitney test. **G.** Venn diagram plotting the intersection of CP with increased level of antibody and T cell response (left) and with increased T cell responses to the S, M and N proteins (right).

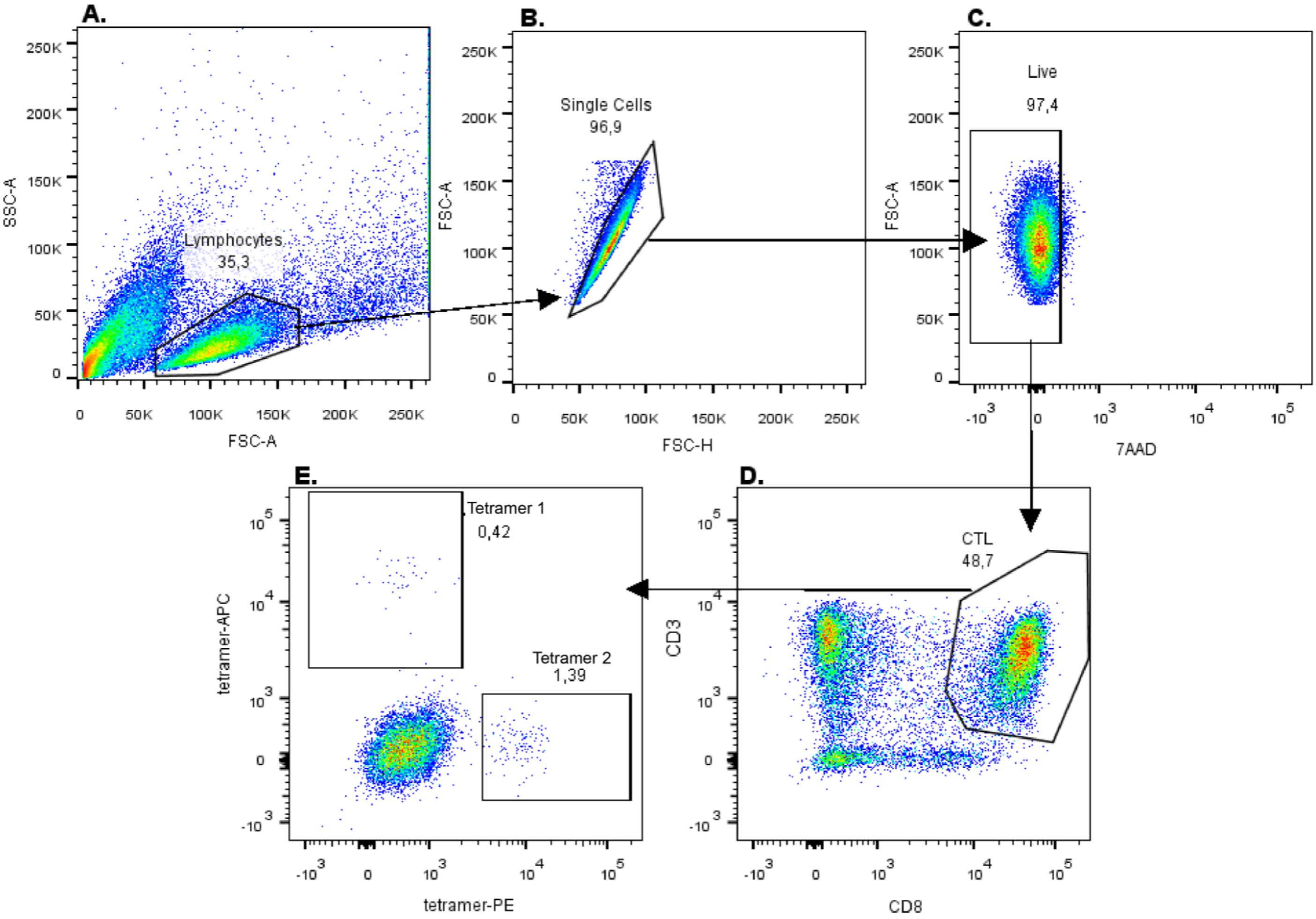


**Figure S3. Flow cytometry gating strategy. A**. Total lymphocytes were gated based on forward scatter (FSC-A)/side scatter (SSC-A). **B.** Singlets were gated based on area and high FSC-A signal. **C.** Live cells were gated based on FSC-A and absence of 7AAD staining. **D.** Cytotoxic T cells were gated based on CD3 and CD8 positivity. **E.** Epitope-specific T cells were gated based on MHC-tetramer-PE or -APC positivity.

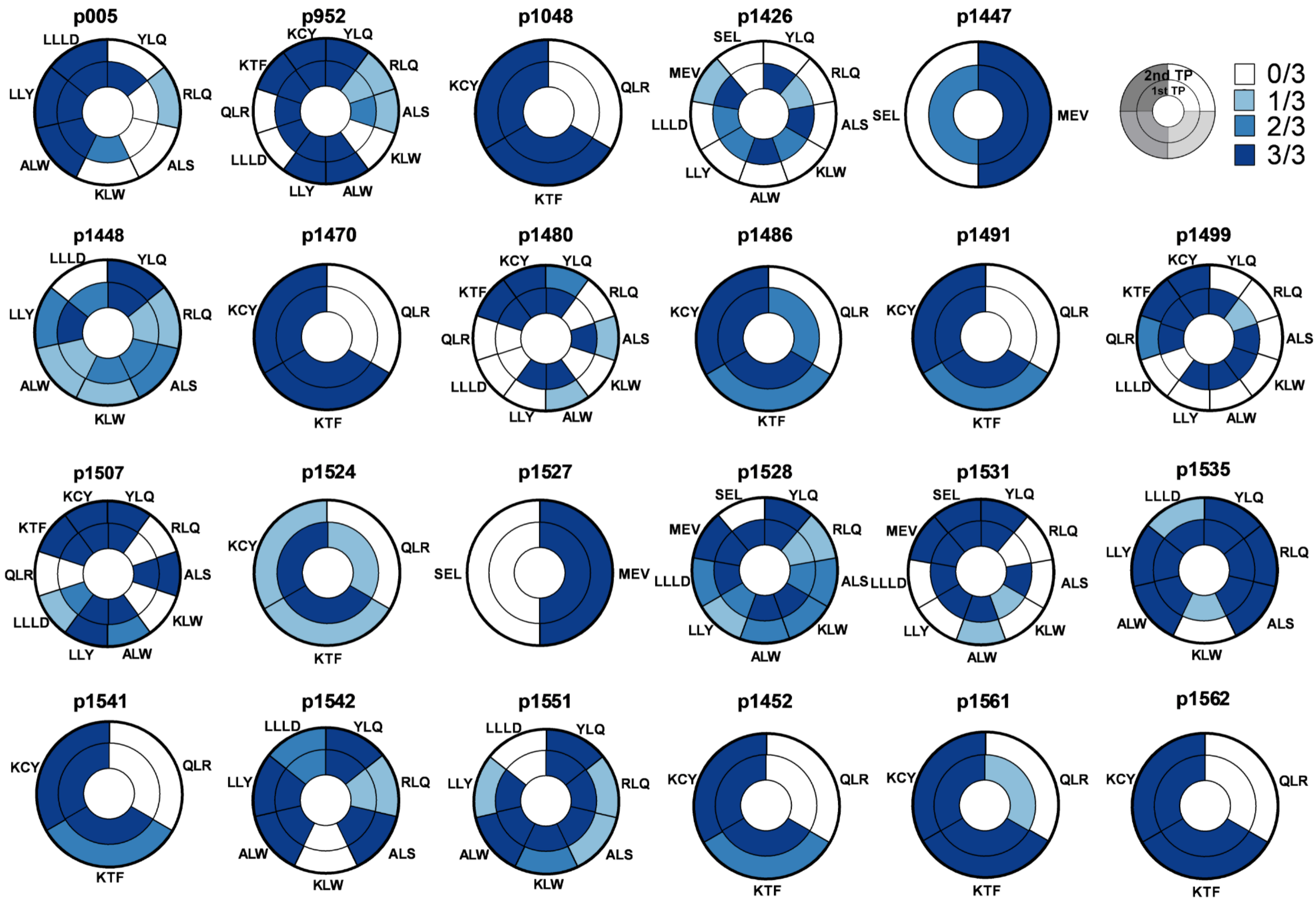


**Figure S4. Change in frequency of antigen-specific cells after rapid in vitro expansion**. Data show relative antigen-specific response at two time-points (TP is inner circle, TP2 is outer circle). Donor ID is indicated on top of each pie chart. Each segment corresponds to one epitope, which is indicated outside by its three- or four-letter code. Color indicates the number of wells with MHC-tetramer$^{+}$ cells.

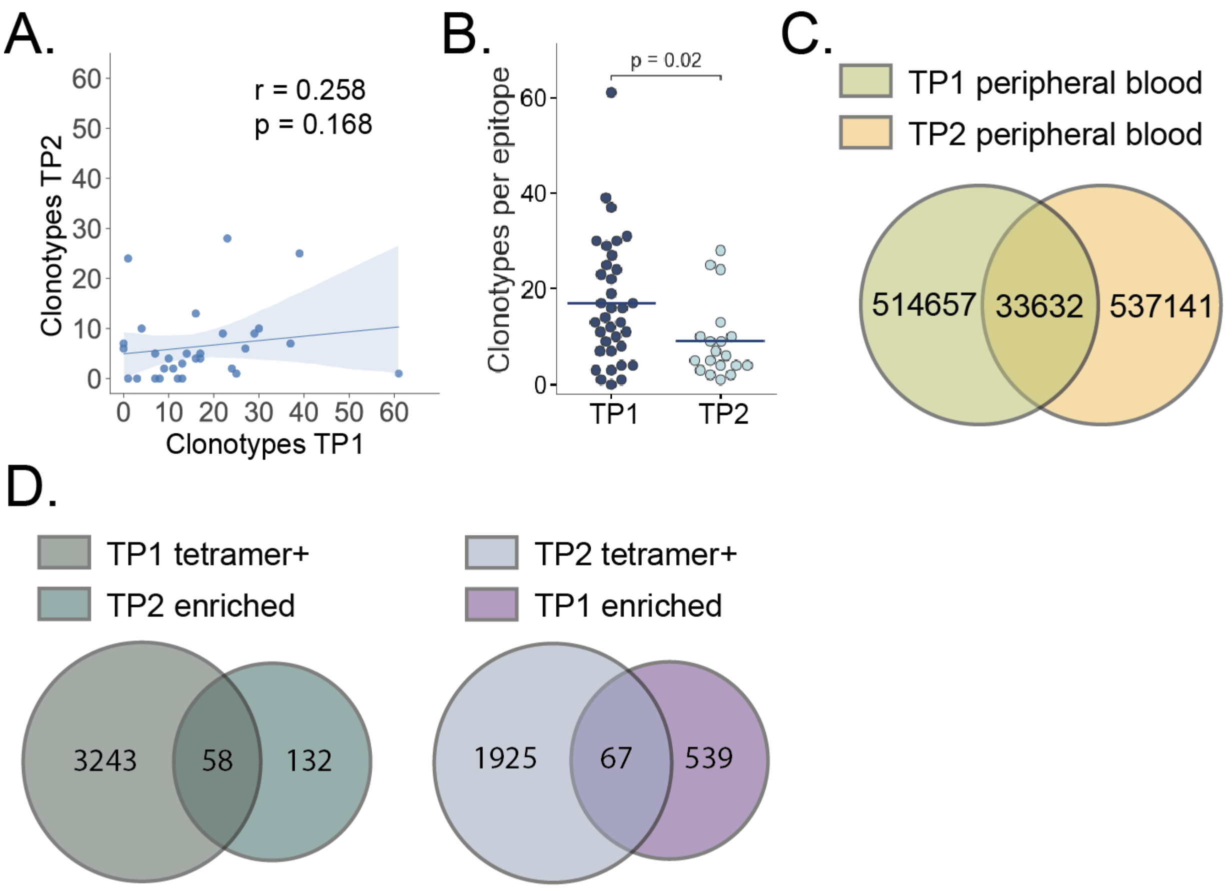


**Figure S5. Correlation between the number of epitopes at different time-points and intersections of different populations. A.** Spearman correlation between the number of clonotypes at TP1 and TP2. r = correlation coefficient. **B.** Number of specific clonotypes for epitopes generating a response in three wells. Mann-Whitney test, statistically significant values are annotated. **C, D.** Venn diagram plotting the intersection of **C,** peripheral blood clones between the two time-points and **D,** all MHC-tetramer$^{+}$ clones and enriched fractions.

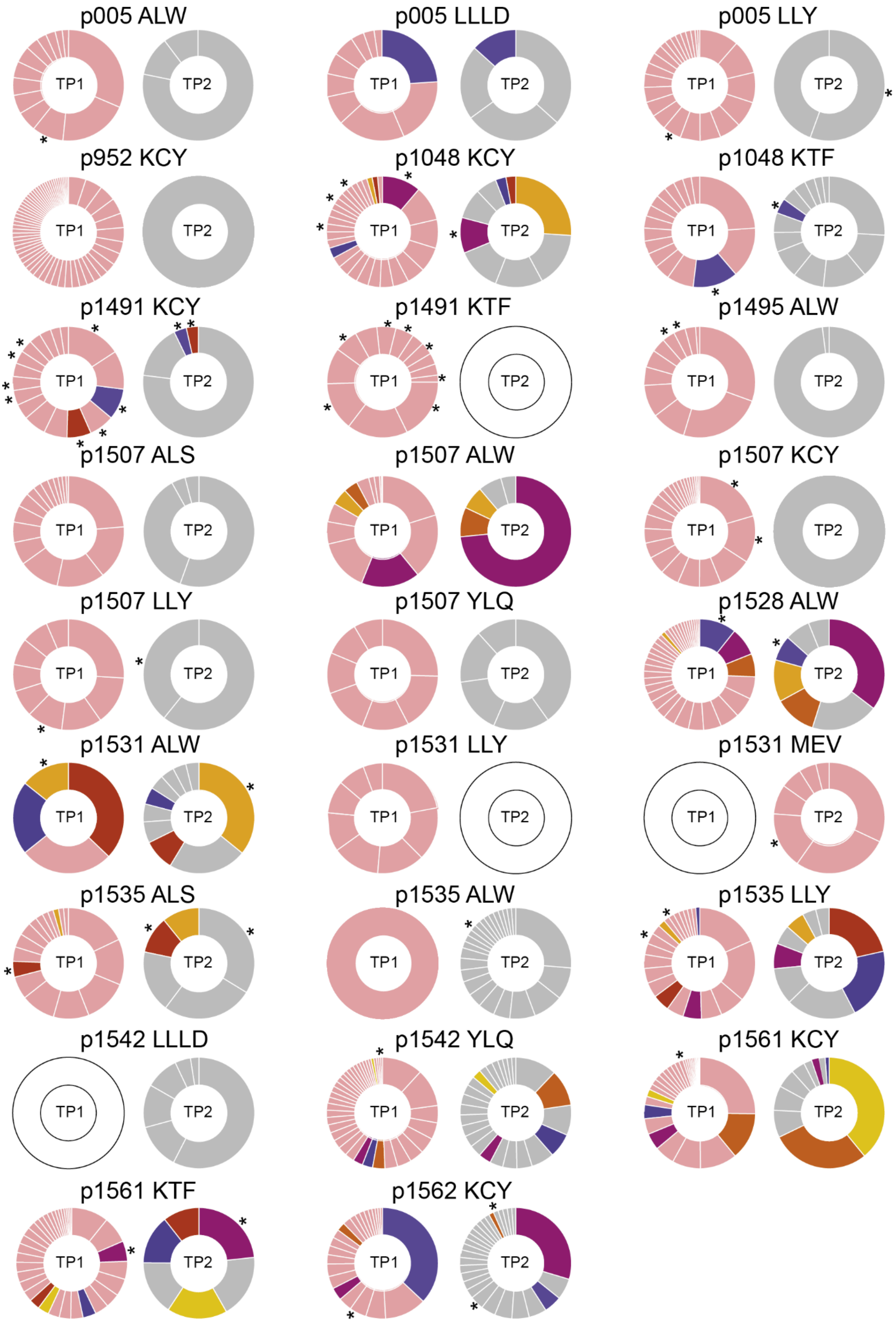
p005 ALW
p005 LLLD
p005 LLY
p952 KCY
p1048 KCY
p1048 KTF
p1491 KCY
p1491 KTF
p1495 ALW
p1507 ALS
p1507 ALW
p1507 KCY
p1507 LLY
p1507 YLQ
p1528 ALW
p1531 ALW
p1531 LLY
p1531 MEV
p1535 ALS
p1535 ALW
p1535 LLY
p1542 LLLD
p1542 YLQ
p1561 KCY
p1561 KTF
p1562 KCY
TP1
TP2

**Figure S6. Clonal structure of $CD8^+$ epitope-specific populations where clones intersecting between the two time-points were found.** Pie charts represent the share of the clonotype in the epitope-specific repertoire at TP1 (left) and TP2 (right). Pink indicates clonotypes found only at TP1, gray indicates clonotypes found only at TP2, other colors indicate clonotypes found at both time-points. Asterisks indicate clonotypes found in total repertoires.

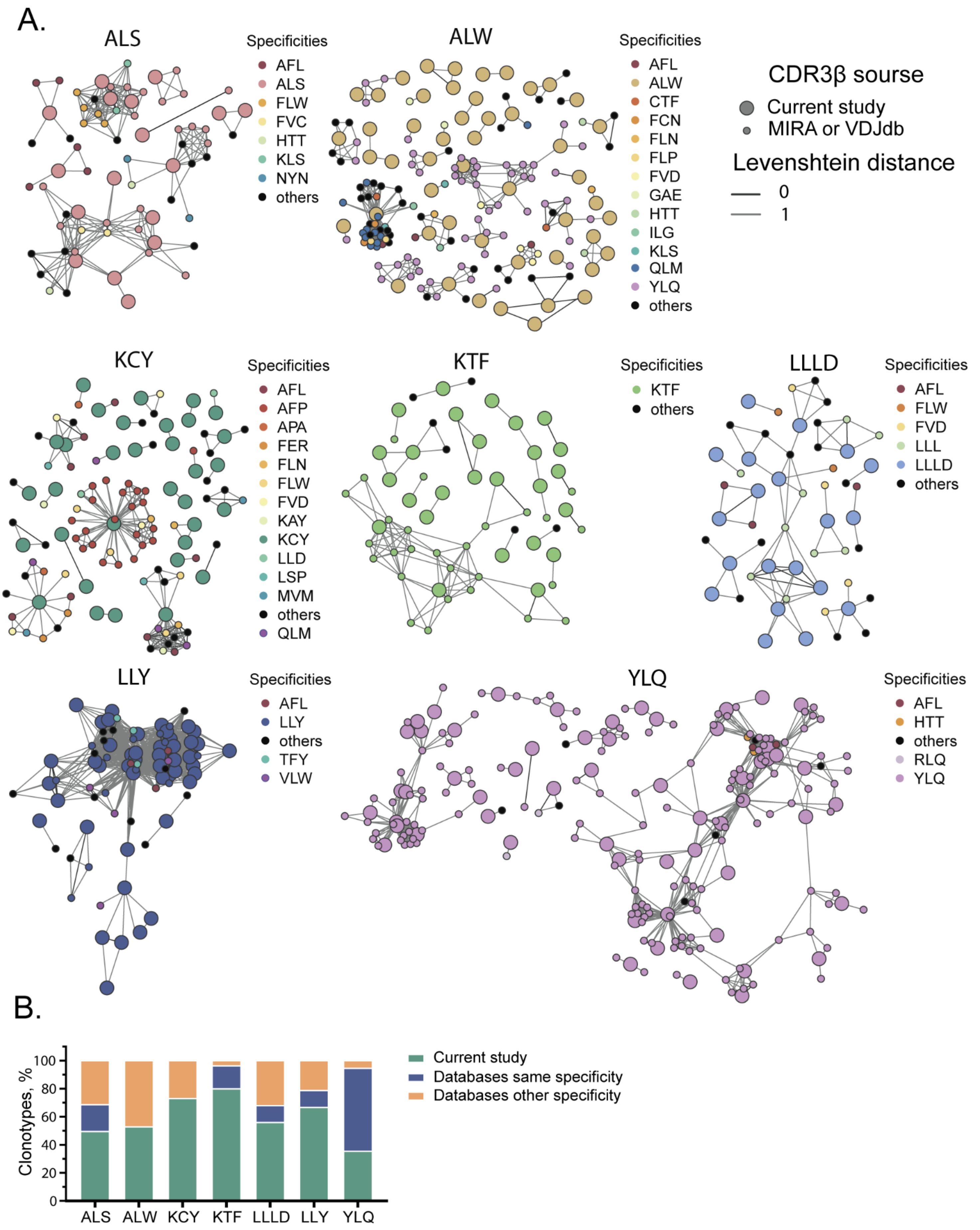

A.
ALS
Specificities
AFL
ALS
FLW
FVC
HTT
KLS
NYN
others
ALW
Specificities
AFL
ALW
CTF
FCN
FLN
FLP
FVD
GAE
HTT
ILG
KLS
QLM
YLQ
others
CDR3β sourse
Current study
MIRA or VDJdb
Levenshtein distance
0
1
KCY
Specificities
AFL
AFP
APA
FER
FLN
FLW
FVD
KAY
KCY
LLD
LSP
MVM
others
QLM
KTF
Specificities
KTF
others
LLLD
Specificities
AFL
FLW
FVD
LLL
LLLD
others
LLY
Specificities
AFL
LLY
others
TFY
VLW
YLQ
Specificities
AFL
HTT
others
RLQ
YLQ
B.
Clonotypes, %
0
20
40
60
80
100
ALS
ALW
KCY
KTF
LLLD
LLY
YLQ
Current study
Databases same specificity
Databases other specificity

**Figure S7. SARS-CoV-2 epitope-specific CDR3β amino acid clonotypes form clusters with different levels of similarity. A.** Nodes representing individual CDR3β sequences. Lines show groups of similar sequences at Levenshtein distance of 1 (grey) or 0 (black). Colors indicate epitope specificities from the current study or from the MIRA and VDJdb databases. Big circles indicate CDR3 from the current study, small circles are from MIRA or VDJdb. Only clusters with two or more members are shown. **B.** Fraction of similar sequences (Levenshtein distance ≤ 1) to CDR3 from the current study (yellow) that are also annotated in the MIRA or VDJdb databases with the same (blue) or differing specificity (green).

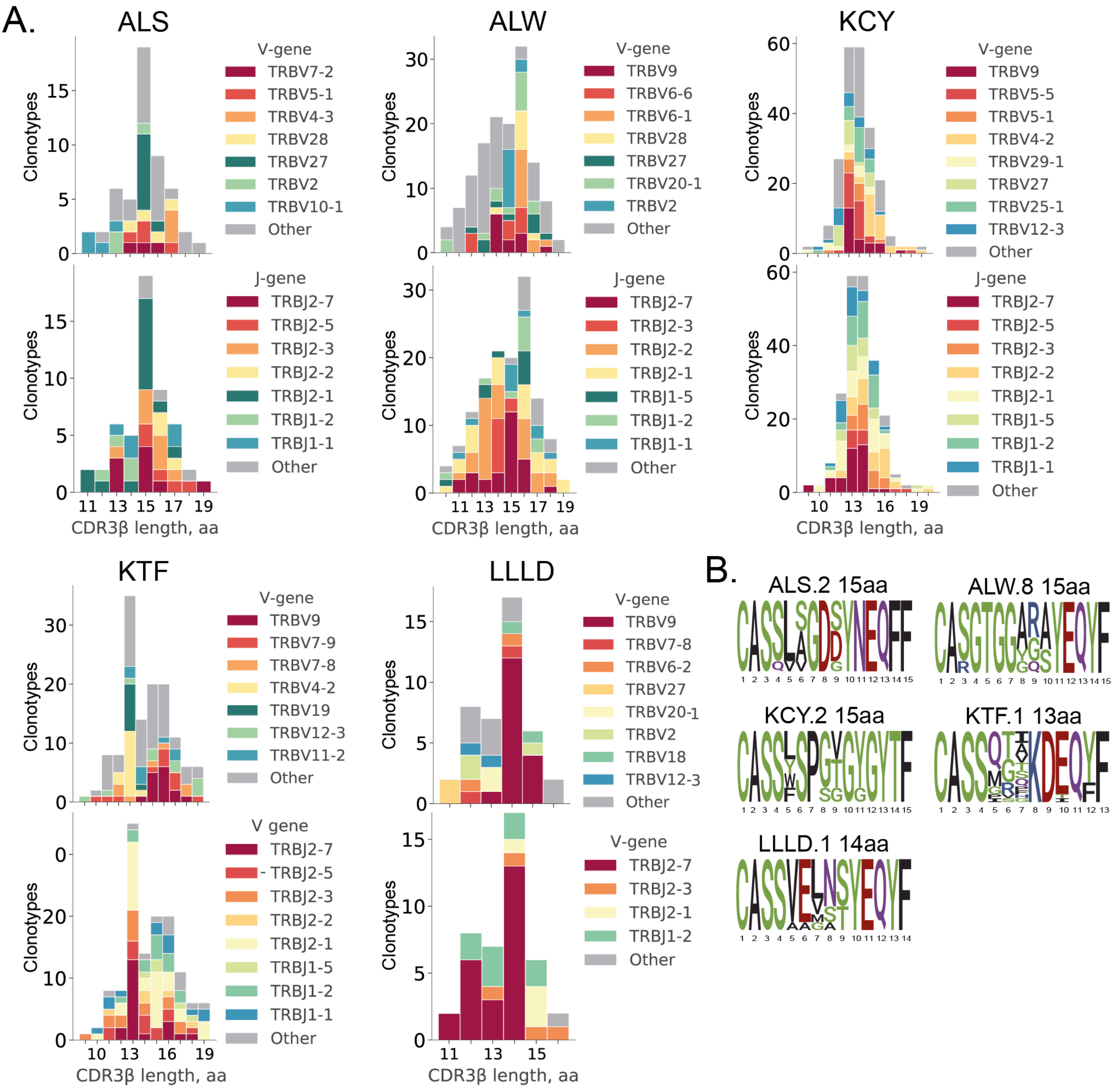

**Figure S8. V and J gene usage of epitope-specific clonotypes and position-weight matrices for CDR3β sequences. A.** Histograms of V (upper plots) and J gene usage (lower plots). **B.** Position-weight matrices for CDR3β sequences with the most common length found in epitope-specific clusters. Cluster numbers correspond to numbers shown in **Fig. 4A.**

**Table S1. Information about convalescent patients (CD) and healthy donors (HD) recruited in study.**

| Group | ID | Sex | Age | TP1 | TP2 | Severity of disease |
|---|---|---|---|---|---|---|
| CP | p1426 | M | 17 | 21 | 209 | Mild |
| CP | p1428 | F | 47 | 25 | 213 | Mild |
| CP | p1436 | F | 29 | 25 | 200 | Mild |
| CP | p1437 | M | 28 | 25 | 251 | Mild |
| CP | p1445 | M | 32 | 33 | 211 | Mild |
| CP | p1446 | F | 36 | 25 | 193 | Mild |
| CP | p1447 | F | 59 | 34 | 204 | Mild |
| CP | p1448 | M | 37 | 30 | 217 | Moderate/severe |
| CP | p1452 | F | 19 | 40 | 208 | Mild |
| CP | p1463 | M | 38 | 35 | 202 | Moderate/severe |
| CP | p1465 | M | 19 | 31 | 210 | Moderate/severe |
| CP | p1466 | M | 38 | 38 | 226 | Mild |
| CP | p1470 | F | 41 | 32 | 200 | Mild |
| CP | p1476 | M | 30 | 34 | 254 | Moderate/severe |
| CP | p1477 | F | 30 | 35 | 220 | Moderate/severe |
| CP | p1480 | M | 29 | 37 | 200 | Moderate/severe |
| CP | p1481 | F | 30 | 49 | 212 | Mild |
| CP | p1482 | F | 36 | 39 | 266 | Moderate/severe |
| CP | p1486 | F | 46 | 42 | 260 | Moderate/severe |
| CP | p1487 | M | 46 | 42 | 260 | Mild |
| CP | p1491 | M | 56 | 17 | 232 | Mild |
| CP | p1495 | M | 41 | 45 | 257 | Mild |
| CP | p1499 | F | 35 | 52 | 263 | Moderate/severe |
| CP | p1507 | M | 29 | 39 | 237 | Mild |
| CP | p1515 | F | 64 | 32 | 235 | Moderate/severe |
| CP | p1521 | F | 36 | 64 | 292 | Mild |
| CP | p1524 | F | 35 | 38 | 235 | Moderate/severe |
| CP | p1526 | M | 50 | 41 | 261 | Mild |
| CP | p1527 | F | 51 | 44 | 264 | Mild |
| CP | p1528 | F | 20 | 41 | 261 | Moderate/severe |

| | | | | | | |
|---|---|---|---|---|---|---|
| CP | p1531 | M | 23 | 24 | 247 | Mild |
| CP | p1532 | F | 56 | 20 | 243 | Moderate/severe |
| CP | p1535 | F | 38 | 52 | 281 | Moderate/severe |
| CP | p1537 | M | 37 | 50 | 271 | Mild |
| CP | p1538 | F | 50 | 34 | 251 | Moderate/severe |
| CP | p1541 | F | 41 | 41 | 256 | Moderate/severe |
| CP | p1542 | M | 40 | 49 | 262 | Mild |
| CP | p1543 | F | 38 | 39 | 251 | Asymptomatic |
| CP | p1550 | F | 58 | 32 | 187 | Mild |
| CP | p1551 | M | 24 | 35 | 246 | Mild |
| CP | p1561 | F | 61 | 63 | 272 | Mild |
| CP | p1562 | M | 30 | 68 | 277 | Mild |
| CP | p1565 | M | 63 | 72 | 281 | Mild |
| CP | p1569 | F | 39 | 35 | 240 | Moderate/severe |
| CP | p1576 | F | 24 | 34 | 243 | Mild |
| CP | p005 | M | 33 | 34 | 213 | Mild |
| CP, HD | p1048 | F | 26 | 25 | 187 | Asymptomatic |
| CP, HD | p952 | M | 36 | 18 | 241 | Mild |
| CP, HD | p006 | M | 24 | 21 | 180 | Mild |
| CP, HD | p859 | F | 25 | 60 | 254 | Mild |
| HD | p846 | F | 24 | N/A | N/A | N/A |
| HD | p1018 | F | 23 | N/A | N/A | N/A |
| HD | p1032 | N/A | N/A | N/A | N/A | N/A |
| HD | p1187 | N/A | N/A | N/A | N/A | N/A |
| HD | p1305 | M | 26 | N/A | N/A | N/A |
| HD | p1440 | F | 30 | N/A | N/A | N/A |
| HD | p818 | F | N/A | N/A | N/A | N/A |
| HD | p1203 | F | N/A | N/A | N/A | N/A |
| HD | p1184 | F | N/A | N/A | N/A | N/A |
| HD | p021 | F | N/A | N/A | N/A | N/A |
| HD | p815 | M | N/A | N/A | N/A | N/A |
| HD | p258 | M | N/A | N/A | N/A | N/A |
| HD | p931 | M | N/A | N/A | N/A | N/A |
| HD | p933 | M | N/A | N/A | N/A | N/A |
| HD | p944 | M | N/A | N/A | N/A | N/A |

CP - convalescent patient, HD - healthy donor, F - female, M - male, N/A - not available

**Table S2. HLA typing of CP and HD used for rapid epitope-specific expansions**

| ID | HLA-A1* | HLA-A1* | HLA-B1* | HLA-B1* | HLA-C1* | HLA-C1* |
|---|---|---|---|---|---|---|
| p1426 | 02:01:01:01 | 24:02:01:01 | 38:01:01:01 | 40:01:02 | 03:04:01:01 | 12:03:01:01 |
| p1445 | 02:01:01:01 | 03:01:01:01 | 44:02:01:01 | 44:27:01:01 | 05:01:01:02 | 07:04:01 |
| p1447 | 31:01:02:01 | 31:01:02:01 | 40:01:02 | 55:02:01:03 | 01:02:01:01 | 03:04:01:01 |
| p1448 | 02:01:01:01 | 26:01:01:01 | 27:05:02:01 | 39:01:01:05 | 07:02:01:03 | 12:03:01:01 |
| p1452 | 03:01:01:01 | 29:01:01:01 | 35:01:01:05 | 44:02:01:01 | 04:01:01 | 16:04:01:01 |
| p1463 | 01:01:01:01 | 32:01:01:01 | 27:05:02:05 | 37:01:01:01 | 01:02:01:01 | 06:02:01:01 |
| p1466 | 11:01:01:01 | 32:01:01:01 | 08:01:01:02 | 52:01:01:02 | 07:02:01:01 | 12:02:02:01 |
| p1470 | 03:01:01:01 | 23:01:01:01 | 35:01:01:05 | 44:03:01:19 | 04:01:01 | 04:09N |
| p1480 | 02:01:01:01 | 03:01:01:03 | 40:01:02 | 58:01:01:03 | 03:02:02:05 | 03:04:01:01 |
| p1482 | 01:01:01:01 | 68:12:01 | 15:01:01:01 | 18:03:01:01 | 06:02:01:01 | 07:01:01 |
| p1486 | 01:01:01:01 | 03:01:01:01 | 35:01:01:05 | 52:01:01:02 | 04:01:01 | 12:02:02:01 |
| p1491 | 03:01:01:01 | 24:02:01:01 | 35:01:01:05 | 35:03:01 | 04:01:01 | 04:01:01 |
| p1495 | 02:01:01G | 23:01:01G | 27:05:02G | 44:03:01G | 02:02:02G | 04:01:01G |
| p1499 | 02:01:01G | 03:01:01G | 07:17 | 51:01:01G | 02:02:02G | 07:02:01G |
| p1507 | 02:01:01G | 03:01:01G | 13:02:01G | 15:01:01G | 03:03:01G | 06:02:01G |
| p1524 | 03:01:01:01 | 26:01:01:01 | 08:01:01:01 | 51:01:01:01 | 07:01:01 | 14:02:01 |
| p1527 | 25:01:01:01 | 25:01:01:01 | 13:02:01:01 | 40:01:02 | 03:04:01:01 | 06:02:01:01 |
| p1528 | 02:01:01:01 | 25:01:01:01 | 18:01:01 | 40:01:02 | 01:02:01:01 | 03:04:01:01 |
| p1531 | 01:01:01:01 | 02:01:01:01 | 15:01:01:01 | 40:01:02 | 03:03:01:01 | 03:04:01:01 |
| p1532 | 02:01:01:01 | 03:01:01:01 | 07:02:01:01 | 40:01:02 | 03:04:01:01 | 07:02:01:03 |
| p1535 | 02:01:01:01 | 02:01:01:01 | 27:05:02:05 | 27:05:02:10 | 01:02:01:01 | 01:02:01:01 |
| p1537 | 01:01:01:01 | 24:02:01:01 | 08:01:01:01 | 37:01:01:01 | 06:02:01:01 | 07:01:01 |
| p1541 | 03:01:01:01 | 68:01:01:02 | 35:03:01 | 51:01:01:10 | 04:01:01 | 05:01:01:02 |
| p1542 | 02:01:01:01 | 02:01:01:01 | 44:02:01:01 | 49:01:01:01 | 07:01:01 | 16:02:01:01 |
| p1550 | 11:01:01:01 | 25:01:01:01 | 15:01:01:01 | 18:01:01 | 04:01:01:05 | 12:03:01:01 |
| p1551 | 02:01:01:01 | 68:01:01:02 | 07:02:01:01 | 44:02:01:01 | 05:01:01:02 | 07:02:01:03 |
| p1561 | 03:01:01:01 | 03:01:01:01 | 07:02:01:01 | 07:02:01:01 | 07:02:01:03 | 07:02:01:03 |
| p1562 | 01:01:01:01 | 03:01:01:01 | 07:02:01:01 | 13:02:01:01 | 06:02:01:01 | 07:02:01:03 |
| p1569 | 02:01:01:01 | 11:01:01:01 | 35:08:01:01 | 57:01:01:01 | 04:01:01:28 | 06:02:01:01 |
| p005 | 1:01 | 2:01 | 8:01:01 | 44 | N/A | N/A |

| | | | | | | |
|---|---|---|---|---|---|---|
| p1048 | 01:01:01:01 | 03:01:01:01 | 41:02 | 52:01:01:02 | 07:01:01 | 17:01 |
| p952 | 2:01 | 03:01 | 49 | 51 | 4 | 7 |
| p859 | 2:01 | 2:01 | 7:02 | 7:02 | 7:02 | 7:02 |
| p1018 | 02:01:01:01 | 1:01 | 35:01:00 | 27:05:00 | 1:02 | 4:01 |
| p818 | 2:01 | 24:02:00 | 40:01:00 | 49:01:00 | 3:04 | 7:01 |
| p1203 | 02:01:01:01 | 03:01:01:01 | 40:01:01 | 40:02:01 | 2:02:02 | 3:04:01 |
| p1184 | 02:01:01:01 | 03:01:01:01 | 13:02:01 | 57:01:01 | 6:02 | 6:02 |
